# Distinctive features of SARS-CoV-2-specific T cells predict recovery from severe COVID-19

**DOI:** 10.1101/2021.01.22.21250054

**Authors:** Jason Neidleman, Xiaoyu Luo, Ashley F. George, Matthew McGregor, Junkai Yang, Cassandra Yun, Victoria Murray, Gurjot Gill, Warner C. Greene, Joshua Vasquez, Sulggi Lee, Eliver Ghosn, Kara Lynch, Nadia R. Roan

## Abstract

Although T cells are likely players in SARS-CoV-2 immunity, little is known about the phenotypic features of SARS-CoV-2-specific T cells associated with recovery from severe COVID-19. We analyzed T cells from longitudinal specimens of 34 COVID-19 patients with severities ranging from mild (outpatient) to critical culminating in death. Relative to patients that succumbed, individuals that recovered from severe COVID-19 harbored elevated and increasing numbers of SARS-CoV-2-specific T cells capable of homeostatic proliferation. In contrast, fatal COVID-19 displayed elevated numbers of SARS-CoV-2-specific regulatory T cells and a time-dependent escalation in activated bystander CXCR4+ T cells. Together with the demonstration of increased proportions of inflammatory CXCR4+ T cells in the lungs of severe COVID-19 patients, these results support a model whereby lung-homing T cells activated through bystander effects contribute to immunopathology, while a robust, non-suppressive SARS-CoV-2-specific T cell response limits pathogenesis and promotes recovery from severe COVID-19.

**Graphical Abstract:** 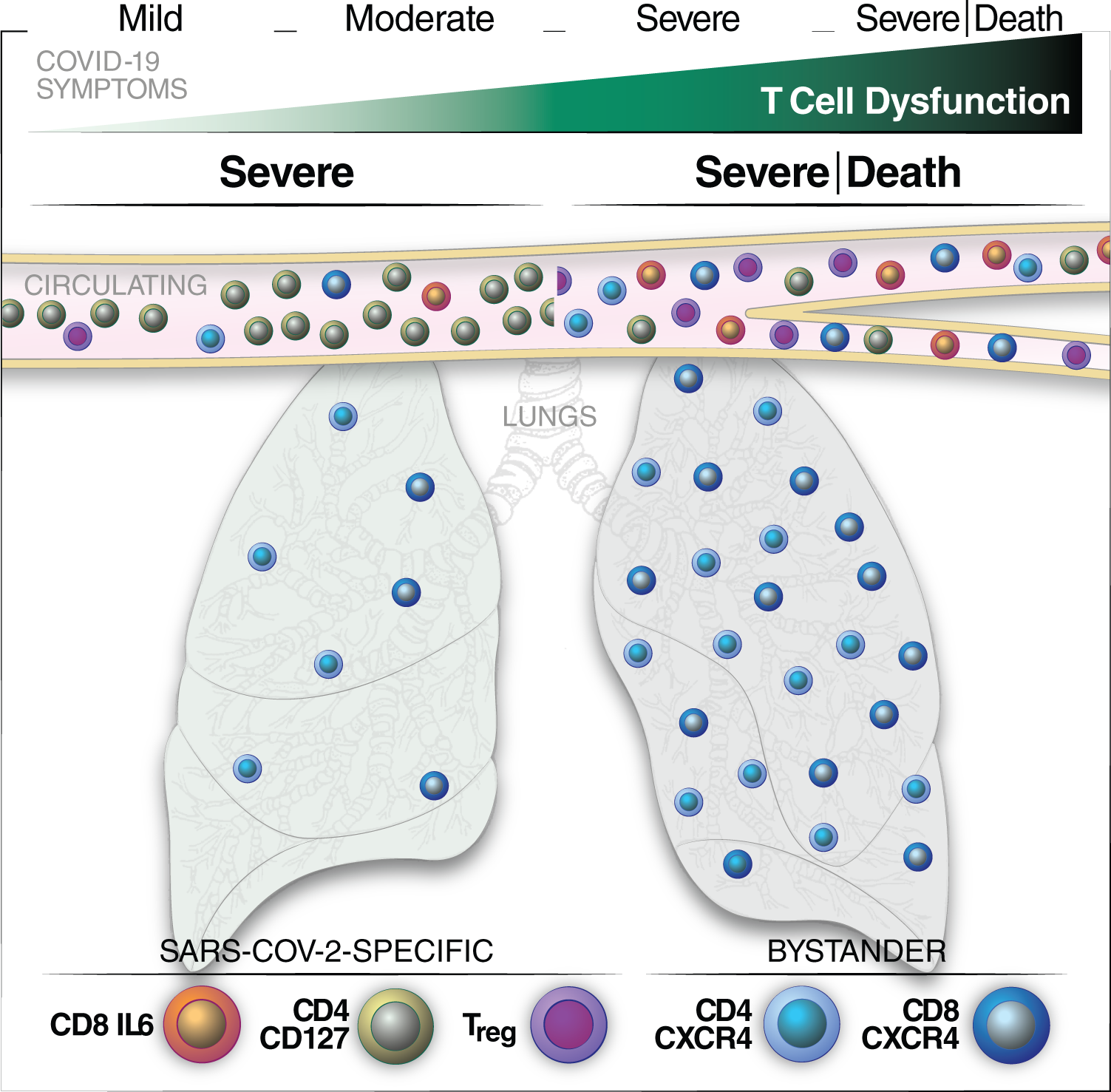

**HIGHLIGHTS:**

- Dysfunctional spike-specific T cells are characteristic of severe COVID-19
- Spike-specific CD127+ Th1 cells are increased in survivors of severe COVID-19
- Spike-specific Tregs and IL6+ CD8+ T cells are increased in fatal COVID-19
- Escalation of activated lung-homing CXCR4+ T cells associates with fatal COVID-19

**BRIEF SUMMARY:** By conducting CyTOF on total and SARS-CoV-2-specific T cells from longitudinal specimens spanning the entire spectrum of COVID-19 diseases, Neidleman et al. demonstrate that spike-specific Th1 cells capable of IL7-dependent homeostatic proliferation predict survival from severe COVID-19, while Tregs and IL6+ CD8+ T cells recognizing spike predict fatal outcome. Fatal COVID-19 is characterized by escalating activation of bystander CXCR4+ T cells in the lungs. Boosting SARS-CoV-2-specific CD4+ T effector responses while diminishing CXCR4-mediated homing may help recovery from severe disease.

## INTRODUCTION

The COVID-19 pandemic caused by the beta-coronavirus SARS-CoV-2 has taken an unprecedented toll on the world’s healthcare systems and economy, and a year since its emergence has already claimed over 2 million lives with fatality rates reaching as high as 20% in some countries (Sorci et al., 2020). While most infected individuals are asymptomatic or mildly symptomatic, up to ∼20% require hospitalization, and this rate increases dramatically in the elderly (>65 years) and those with underlying health conditions (Docherty et al., 2020; Grasselli et al., 2020). Evidence to date suggests variability in host response, rather than viral genetics, to be the prime driver behind the wide range of disease manifestation. For example, individuals genetically pre-disposed to low type I IFN activity, due to inborn loss-of-function genetic variants or autoantibodies against these innate immune cytokines, have increased risk of severe disease (Bastard et al., 2020; van der Made et al., 2020; Zhang et al., 2020).

The adaptive immune system, consisting of cellular (T cell) and humoral (B cell) immunity, is also important in the host’s defense against SARS-CoV-2. While a coordinated response between the cellular and humoral arms seems to be important in effective control (Rydyznski Moderbacher et al., 2020), T cells appear able to resolve infection when B cell responses are insufficient. Indeed, the recovery of two individuals with X-linked agammaglobulinemia without the need for oxygen supplementation or intensive care suggests that antibodies are not absolutely required for clearing SARS-CoV-2 (Soresina et al., 2020). In fact, high levels of anti-SARS-CoV-2 antibodies are associated with more severe disease (Garcia-Beltran et al., 2020; Liu et al., 2019; Woodruff et al., 2020), suggesting that high antibody levels may not always be effective. A consistent hallmark of severe COVID-19 is T cell lymphopenia (Chen et al., 2020a; Zhao et al., 2020a), which is unlikely to simply reflect T cell sequestration in the infected lungs (Liao et al., 2020). Importantly, while overall T cell lymphopenia is observed, the frequencies of some T cell subsets positively associate with disease severity. For example, activated T cells and regulatory T cells (Tregs) have been reported to be elevated in severe cases (De Biasi et al., 2020; Mathew et al., 2020). It is however unclear whether these T cells are specific or not to SARS-CoV-2.

Indeed, while many studies have profiled total T cells across the entire spectrum of COVID-19 severity, few studies have analyzed the features of T cells recognizing SARS-CoV-2 epitopes. As these antigen-specific cells are the ones that can directly recognize virally-infected cells and aid in the generation of SARS-CoV-2-specific antibodies, they have the most potential to exert a beneficial effect on recovery from disease and are the T cell targets of vaccination.

We recently demonstrated that SARS-CoV-2-specific T cells from convalescent individuals that had recovered from mild disease produced IFNγ, but not IL4, IL6, or IL17 (Neidleman et al., 2020a). The undetected cytokines, particularly IL6, have been implicated in COVID-19-associated pathogenesis (Del Valle et al., 2020; Hotez et al., 2020; Huang et al., 2020; Mathew et al., 2020; Pacha et al., 2020; Zhou et al., 2020), although whether SARS-CoV-2-specific T cells secrete these cytokines during severe disease, and, if so, whether this contributes to pathogenesis, are not clear. In fact, the fundamental question of whether SARS-CoV-2-specific T cells are beneficial or detrimental during acute infection is debated. A recent study found that a robust SARS-CoV-2-specific T cells, but not a strong antibody response, negatively associates with disease severity (Mathew et al., 2020), while another research group reported a pathogenic role for SARS-CoV-2-specific T cells (Anft et al., 2020; Thieme et al., 2020).

Differences in experimental design may account for these discrepant results, but, importantly, these prior studies did not compare fatal to non-fatal COVID-19 cases. Most studies to date have compared immune features associated with moderate versus severe COVID-19 cases, and to our knowledge no studies have compared the phenotypes of SARS-CoV-2-specific T cells in severe cases of COVID-19 that ultimately recover from or succumb to disease.

Here, we define the features T cells from individuals hospitalized in the ICU for COVID-19, including some that recovered and some that died from disease. We implemented deep-phenotyping of both total and SARS-CoV-2-specific T cells using 38-parameter CyTOF, a technique we recently applied to characterize T cells from convalescent individuals (Neidleman et al., 2020a). By conducting longitudinal assessments on 34 COVID-19 patients with disease manifestations ranging from mild to fatal, we identified unique phenotypic features of SARS-CoV-2-specific and bystander T cells associated with recovery from severe disease, and paired this analysis with a detailed examination of the features of T cells in the lungs of COVID-19 patients.

## RESULTS

### Patient cohorts and experimental design

The demographics and clinical features of the COVID-19 patient cohort are presented in Table S1. We analyzed a total of 48 blood specimens from 34 SARS-CoV-2-infected individuals, along with samples from 11 uninfected controls. Of the 48 specimens, 33 were from individuals hospitalized in the ICU (“severe”) and 6 from patients hospitalized but not in the ICU (“moderate”). The last 9 came from patients who were never hospitalized (“mild”). An additional 11 SARS-CoV-2-negative individuals hospitalized in the ICU were phenotyped by CyTOF. In total, 59 specimens were analyzed for total and SARS-CoV-2-specific T cells. All specimens were analyzed fresh, on the day of the blood draw, to avoid potential confounders associated with cell cryopreservation. Immediately following PBMC isolation, a portion of the cells was fixed for CyTOF, while remaining cells were stimulated with overlapping peptides from SARS-CoV-2 spike to enable detection of induced cytokines. The unstimulated (“baseline”) and peptide-stimulated specimens were analyzed simultaneously using the CyTOF panel presented in Table S2. This panel was designed to include multiple markers of T cell differentiation state, activation status, exhaustion, proliferative potential, and homing properties.

### SARS-CoV-2-specific CD4+ and CD8+ T cells are elicited during mild, moderate, and severe COVID-19

Manual gating was used to identify CD4+ and CD8+ T cells from the 59 specimens analyzed by CyTOF. Because activation markers but not IFNγ were expressed in the baseline specimens (Fig. S1A, Fig. 1A), we used IFNγ to identify SARS-CoV-2-specific T cells in the stimulated samples, similar to our recent study (Neidleman et al., 2020a). While cells producing high levels of IFNγ were readily identified following peptide stimulation (Fig. 1A), cells producing high levels of cytokines IL4, IL6, or IL17 were not observed (Fig. S1B, C). IFNγ+ cells were detected in the stimulated samples from all three patient groups and amounted to a total of 3,735 cells. Their frequencies ranged from undetectable to almost 2% of the patients’ T cells, and were not significantly different between the groups (Fig. 1B). In comparison, total (non-antigen-specific) CD4+ and CD8+ T cell frequencies from the baseline specimens were significantly diminished in the moderate and severe groups (Fig. 1C), consistent with the lymphopenia previously reported in hospitalized, acutely infected individuals (Chen et al., 2020a; Zhao et al., 2020a).

**Figure 1.**
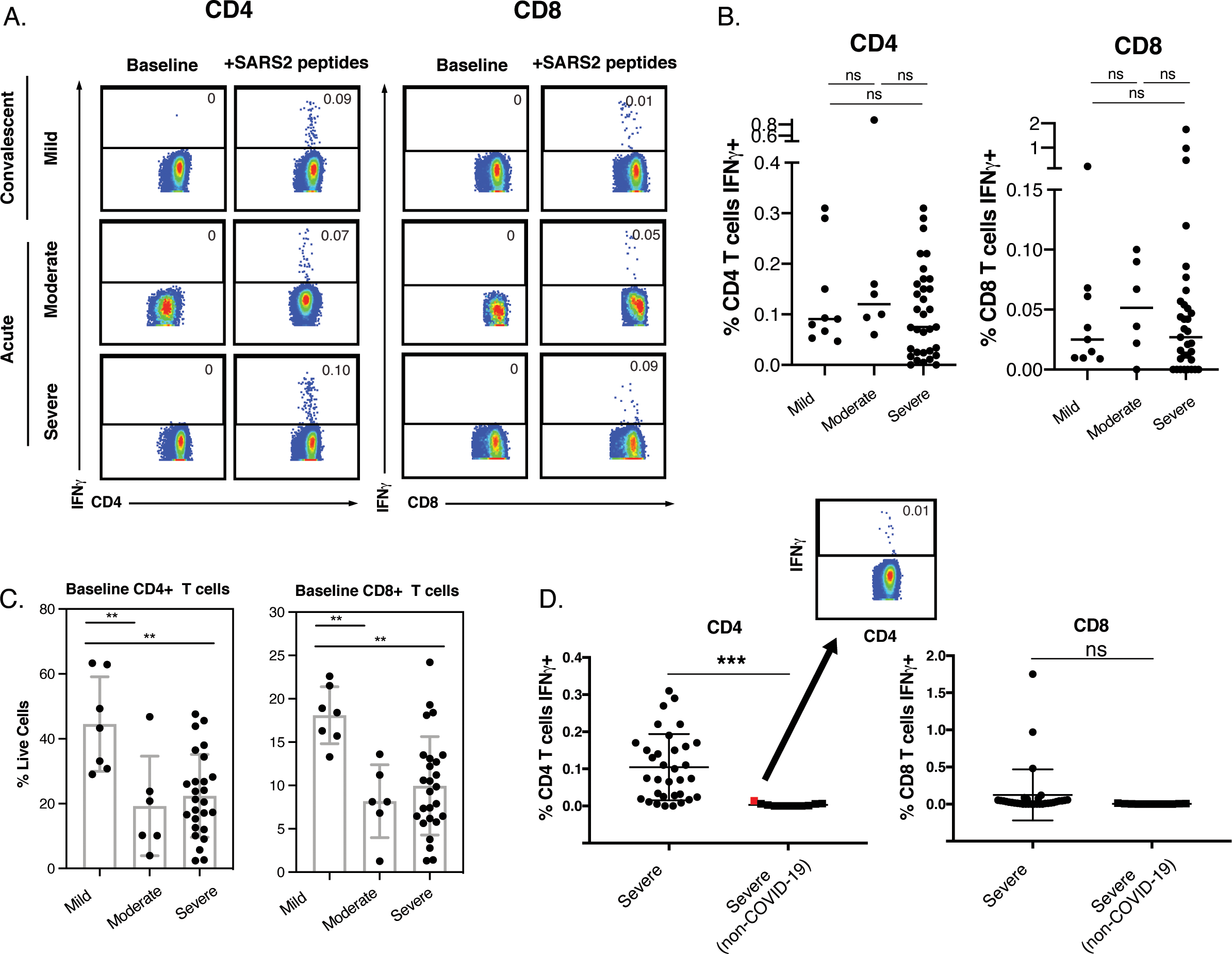
De novo IFNγ-producing SARS-CoV-2 spike-specific CD4+ and CD8+ T cells are elicited during mild, moderate, and severe COVID-19. (A) Pseudocolor plots of CyTOF datasets reflecting percentages of CD4+ or CD8+ T cells producing IFNγ at baseline or in response to a 6-hour stimulation with overlapping peptides from SARS-CoV-2 spike in representative donors from mild, moderate, and severe cases. Results are gated on live, singlet CD4+ or CD8+ T cells. (B) Percentages of CD4+ and CD8+ T cells producing IFNγ in response to spike peptide stimulation in all COVID-19 cases. ns: non-significant as determined by one-way analysis of variance with a Bonferroni post-test. (C) Evidence for CD4+ and CD8+ T cell lymphopenia in specimens from moderate and severe COVID-19 relative to mild cases. ** p < 0.01 as determined by one-way analysis of variance with a Bonferroni post-test. (D) Proportion of IFNγ-producing cells among T cells from ICU patients with or without SARS-CoV-2 infection. Even in the uninfected specimen with the highest response to spike stimulation (*red dot*), the proportion of IFNγ-producing cells was only 0.01% (*inset*), suggesting that the responses we detect in COVID-19 specimens correspond to *de novo* SARS-CoV-2-specific T cells. *** p < 0.001 as determined by a Student’s unpaired t-test. See also Fig. S1, S2 and S3.

As cross-reactive T cell responses have been reported in some individuals never exposed to SARS-CoV-2 (Braun et al., 2020; Grifoni et al., 2020; Sekine et al., 2020), we assessed to what extent the responses we detected in COVID-19 patients were generated *de novo*. We acquired 11 fresh blood specimens from individuals hospitalized in the ICU for reasons unrelated to COVID-19 and confirmed by RT-PCR to not be infected with SARS-CoV-2. SARS-CoV-2-specific CD4+ T cells were significantly more frequent in the infected ICU patients than the uninfected ICU patients (Fig. 1D), confirming *de novo* production of SARS-CoV-2-specific CD4+ T cells by severe COVID-19 patients. Although we did see evidence of low levels of cross-reactive responses, even in the uninfected individual with the strongest response to peptide stimulation, SARS-CoV-2-specific cells only accounted for 0.01% of the CD4+ T cells (Fig. 1D, inset). SARS-CoV-2-specific CD8+ T cells recognizing spike were also detected at higher frequencies in the infected relative to uninfected individuals. However, due to their low frequencies relative to CD4+ T cells (consistent with prior observations (Ferretti et al., 2020; Neidleman et al., 2020a)), this did not reach statistical significance (Fig. 1D). Together, these data suggest that despite lymphopenia, *de novo* T cell responses are mounted against SARS-CoV-2 in patients hospitalized for COVID-19.

### Phenotypes of total and SARS-CoV-2-specific T cells differ in mild, moderate, and severe COVID-19

As an initial examination of the phenotypes of the T cells in infected patients, we calculated the mean signal intensity (MSI) of each antigen (Fig. S2, Fig. S3), and compared them between the 3 patient groups for the following cell populations: Baseline, Bystander, and SARS-CoV-2-specific. Among baseline samples, CD4+ and CD8+ T cells from severe cases expressed significantly higher levels of activation markers HLADR, CD69, CD25, and PD1, as well as chemokine receptor CXCR4, which directs cells to inflamed and damaged lung tissues (Mamazhakypov et al., 2019), relative to mild cases. Among SARS-CoV-2-specific T cells, antigens significantly upregulated in severe relative to mild cases included the activation/exhaustion markers PD1 and TIGIT.

We next assessed the subset distribution of total and SARS-CoV-2-specific T cells using manual gating. CD45RO expression is commonly used to define memory T cells, and CD45RA to define naïve T cells (Tn), although CD45RA is also expressed on terminally differentiated effector memory (Temra) and stem cell memory (Tscm) T cells. We found that for both CD4+ and CD8+ T cells, CD45RO+CD45RA-frequencies were highest in the severe group while CD45RO-CD45RA+ were the lowest (Fig. 2A, B). This was observed for both total and SARS-CoV-2-specific T cells, and is consistent with the severe group expressing overall higher levels of CD45RO and lower levels of CD45RA (Fig. S2, S3). Gating within the CD45RO+CD45RA-and CD45RO-CD45RA+ populations allowed us to identify the proportions of central memory (Tcm), effector memory (Tem), transitional memory (Ttm), Tn, Temra, and Tscm subsets (Fig. 2A, B). SARS-CoV-2-specific CD4+ T cells were enriched for Tem cells and relatively depleted of Temra cells. Changes in subset distribution were also observed among total T cells. For example, the severe group harbored significantly higher proportions of CD8+ Tem and CD4+ Tscm, and lower proportions of CD8+ Ttm.

**Figure 2.**
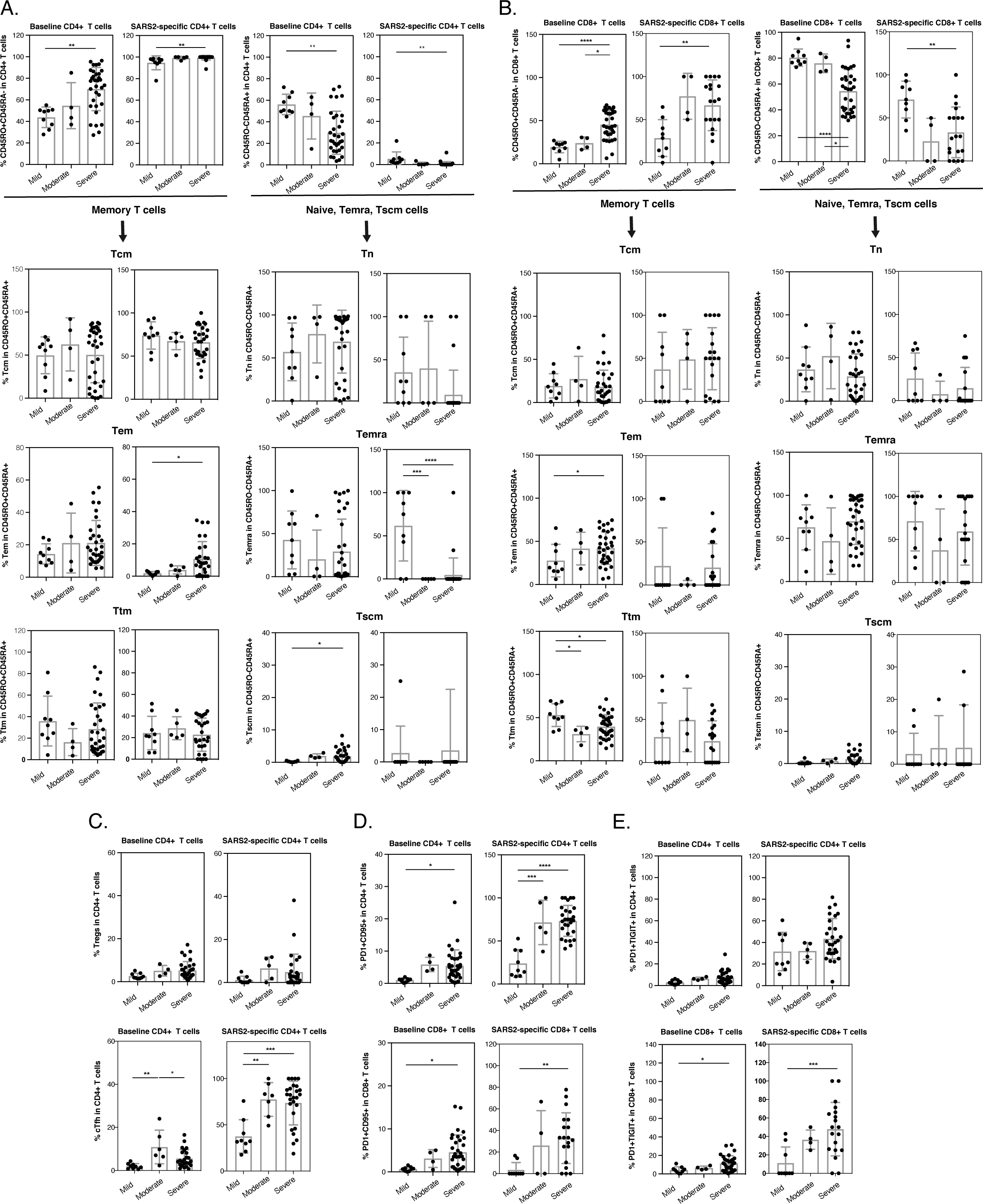
Subset distribution of total and SARS-CoV-2-specific T cells during mild, moderate, and severe COVID-19 by manual gating. (**A, B**) Distribution of baseline and SARS-CoV-2-specific CD4+ (*A*) and CD8+ (*B*) T cells from COVID-19 patients among memory (CD45RO+CD45RA-) and a mixture of Tn, Temra, and Tscm (CD45RO-CD45RA+) cells. Breakdown into further subsets was achieved by manually gating on subset marker combinations. (**C**) Proportions of Tregs (CD45RO+CD45RA-CD25+CD125^low^) and cTfh (PD1+CXCR5+) cells among baseline and SARS-CoV-2-specific CD4+ T cells from COVID-19 patients. (**D**) The proportion of apoptosis-prone (PD1+CD95+) cells among CD4+ and CD8+ T cells is increased in severe relative to mild COVID-19 cases for both total and SARS-CoV-2-specific T cells. (**E**) The proportion of exhausted cells (PD1+TIGIT+) among total and SARS-CoV-2-specific CD8+ T is increased in severe relative to mild COVID-19. * p < 0.05, ** p < 0.01, *** p < 0.001, **** p < 0.0001 as determined by one-way analysis of variance with a Bonferroni post-test.

We then examined the percentages of CD4+ Tregs and circulating T follicular helper (cTfh) cells, subsets important in immunoregulation and helping antibody responses, respectively. The frequencies of Tregs were similar between the three patient groups, although there was a trend for higher levels in the severe group. By contrast, the moderate and severe groups had significantly higher frequencies of SARS-CoV-2-specific cTfh than the mild group did (Fig. 2C), suggesting potential elicitation of antibody production through T-cell help during moderate/severe acute infection. Additional manual gating revealed PD1+CD95+ T cells, within both the total and SARS-CoV-2-specific compartments, to be most abundant during severe infection (Fig. 2D). Total and SARS-CoV-2-specific PD1+TIGIT+ T cells were also more frequent during severe infection, but only within the CD8 compartment (Fig. 2E). These findings are consistent with the increased expression of PD1, CD95, and TIGIT in severe cases (Fig. S2, S3). As co-expression of PD1 and CD95 is characteristic of apoptosis-prone cells while co-expression of PD1 and TIGIT is characteristic of exhausted cells, these findings together suggest an increased T cell dysfunction during severe COVID-19.

### Clustering reveals an accumulation in activation and exhaustion markers in T cells from severe COVID-19 patients

Having analyzed cellular subsets through manual gating, we then implemented a more holistic approach integrating all the CyTOF phenotyping markers in our panel to identify populations of cells associated with disease severity. Clustering by FlowSOM (Van Gassen et al., 2015) revealed differences between mild, moderate, and severe infection in both total and SARS-CoV-2-specific T cells, and among both the CD4+ (Fig. 3A-D) and CD8+ (Fig. 4A-D) compartments. Among total CD4+ T cells, the most noticeable difference was a single cluster (B4.9), composed of activated memory CD4+ T cells expressing high levels of the homing receptor CCR6 (Ito et al., 2011), which was present at higher frequencies in the severe group. However, this difference became insignificant after correction for multiple testing (Fig. 3E). In comparison, total CD8+ T cells revealed four clusters that were significantly enriched in the severe group (Fig. 4E). These clusters all expressed elevated levels of HLADR, but only some expressed CD38 (Fig. 4F), suggesting that HLADR may be a more universal marker of SARS-CoV-2-induced global T cell activation than CD38. One cluster (B8.1) was enriched in the mild group (Fig. 4E), albeit insignificantly after adjustment for multiple correction, and this cluster expressed low levels of both HLADR and CD38 (Fig. 4F).

**Figure 3.**
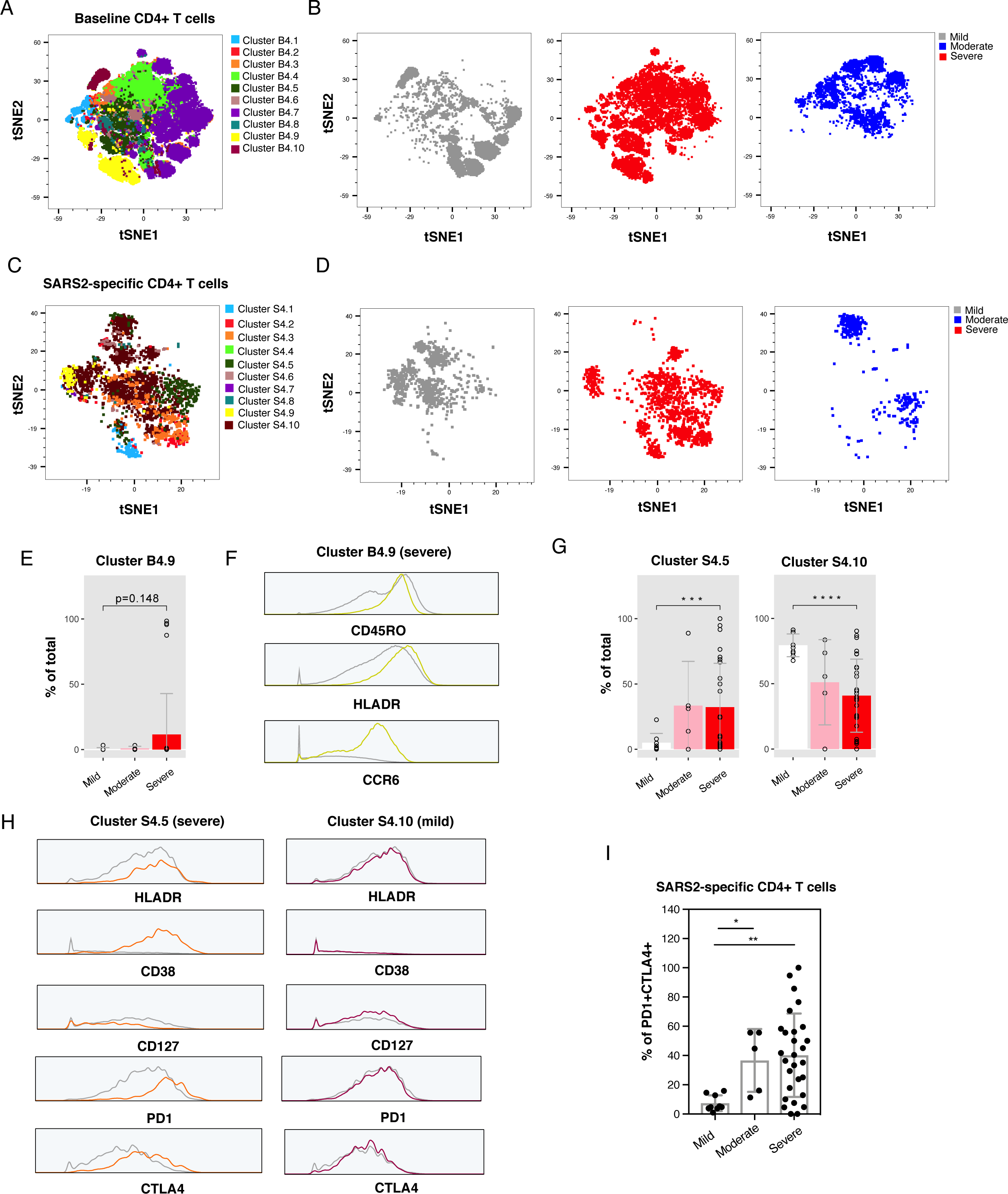
Clustering reveals enrichment in activated and exhausted SARS-CoV-2-specific CD4+ T cells during severe COVID-19. (**A, C**) FlowSOM clustering of total (*Baseline, A*) and SARS-CoV-2-specific (*C*) CD4+ T cells from all infected patient groups, visualized as a t-SNE plot. Ten clusters were identified. Each cluster name begins with a “B” (for <u>b</u>aseline) or an “S” (for <u>S</u>ARS-CoV-2-specific), followed by a “4” (for CD<u>4</u>). (**B, D**) Total (*B*) and SARS-CoV-2-specific (*D*) CD4+ T cells from mild (*grey*), moderate (*blue*), and severe (*red*) COVID-19 map to different areas of the *panel A* t-SNE plot, indicating that they have different phenotypes. (**E**) Proportion of total CD4+ T cells in the B4.9 cluster in the three patient groups. B4.9 enrichment in the severe relative to the mild group was significant (p < 0.05) prior to adjustment for multiple testing by Bonferroni. (**F**) High expression levels of memory marker CD45RO, activation marker HLADR, and chemokine receptor CCR6 in cells from the B4.9 cluster (*yellow trace*) relative to total CD4+ T cells (*grey trace*). (**G**) Proportion of SARS-CoV-2-specific CD4+ T cells in the S4.5 and S4.10 clusters across the three patient groups. Cluster S4.5 was significantly enriched in the severe relative to the mild group, and cluster S4.10 in the mild relative to the severe group. *** p < 0.001 and **** p < 0.0001 as determined by Student’s unpaired t-testand adjusted for multiple testing using Bonferroni for FDR. (**H**) Cluster S4.5 cells express elevated levels of the activation markers HLADR and CD38 and the exhaustion markers PD1 and CTLA4, but diminished levels of the IL7 receptor alpha chain CD127 (*orange traces*) relative to all SARS-CoV-2-specific CD4+ T cells (*grey traces*). Cluster S4.10 cells (*maroon traces*) do not. (**I**) Increased frequencies of exhausted (PD1+CTLA4+) cells among SARS-CoV-2-specific CD4+ T cells in severe relative to mild COVID-19 samples, as validated by manual gating. * p < 0.05 and ** p < 0.01 as determined by one-way analysis of variance with a Bonferroni post-test.

**Figure 4.**
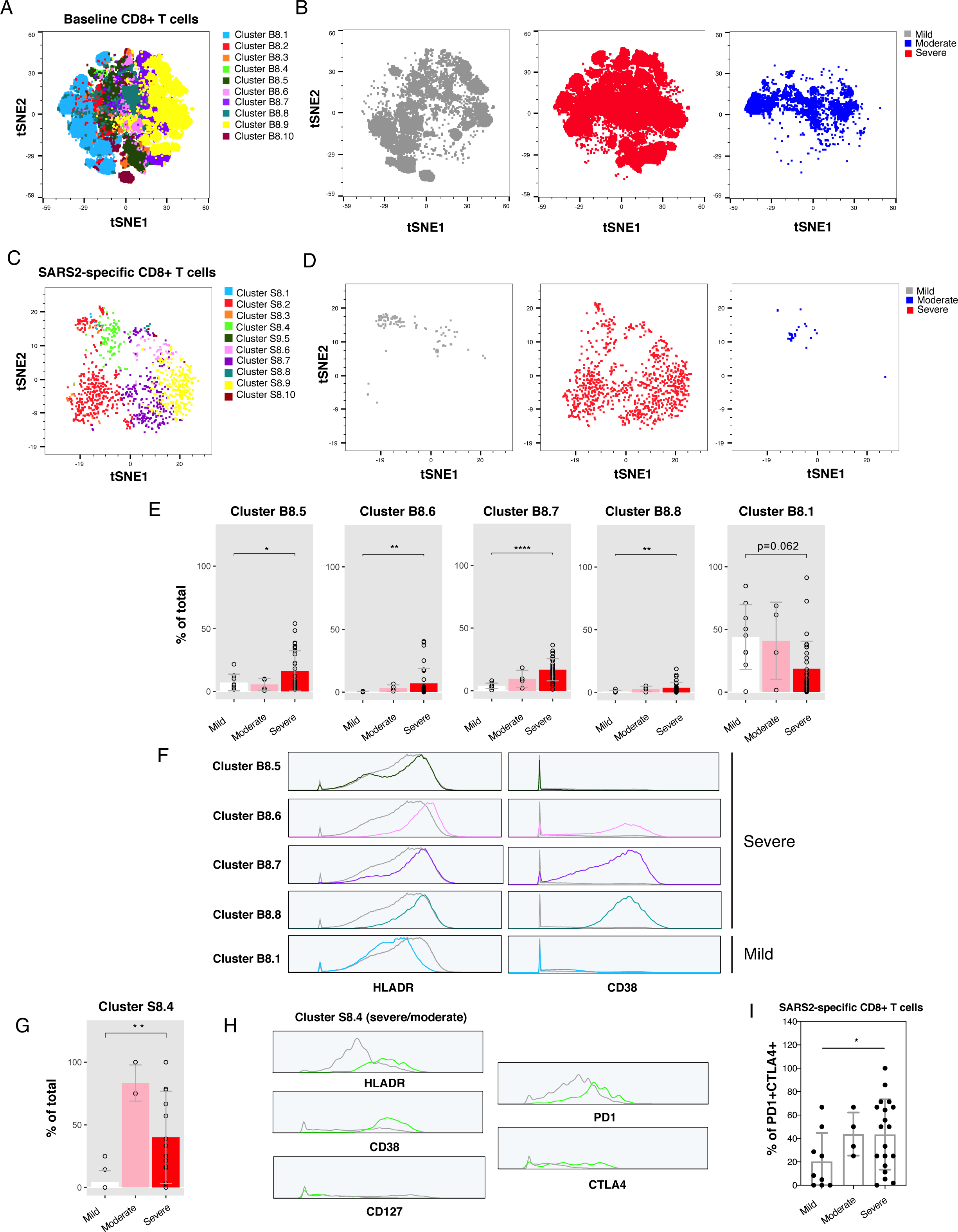
Clustering reveals enrichment of activated, exhausted SARS-CoV-2-specific CD8+ T cells during severe COVID-19. (**A, C**) FlowSOM clustering of total (*Baseline, A*) and SARS-CoV-2-specific (*C*) CD8+ T cells from all infected patient groups, visualized as t-SNE plots. Ten clusters were identified. Each cluster name begins with a “B” (for <u>b</u>aseline) or an “S” (for <u>S</u>ARS-CoV-2-specific), followed by a “4” (for CD<u>4</u>). (**B, D**) Total (*B*) and SARS-CoV-2-specific (*D*) CD8+ T cells from mild (*grey*), moderate (*blue*), and severe (*red*) COVID-19 map to different areas of the t-SNE plots, indicating that they have different phenotypes. (**E**) Clusters B8.5, B8.6, B8.7 and B8.8 of total CD8+ T cells are significantly enriched in severe relative to mild group, while there is a trend for cluster B8.1 enrichment in mild relative to severe group. * p < 0.05, ** p < 0.01, and **** p < 0.0001 as determined by Student’s unpaired t-testand adjusted for multiple testing using Bonferroni for FDR. Of note, when not adjusting for multiple testing, cluster B8.1 was significantly enriched (p < 0.05) in mild vs. severe groups. (**F**) Cells from the B8.5, B8.6, B8.7 and B8.8 (severe-enriched) clusters all express high levels of the activation marker HLADR (*colored traces*) relative to total CD8+ T cells (*grey trace*), while activation marker CD38 is elevated in only a subset of these clusters (*colored traces*) relative to total CD8+ T cells (*grey trace*). Cells from the B8.1 (mild-enriched) cluster express low levels of both activation markers. (**G**) Cluster S8.4 is enriched in the severe and moderate groups relative to the mild group. ** p < 0.01 as determined as assessed by Student’s unpaired t-testand adjusted for multiple testing using Bonferroni for FDR. (**H**) Cluster S8.4 cells express elevated levels of the activation markers HLADR and CD38 and the exhaustion markers PD1 and CTLA4, but diminished levels of the IL7 receptor alpha chain CD127 (*green traces*) relative to all SARS-CoV-2-specific CD8+ T cells (*grey traces*). (**I**) Increased frequencies of exhausted (PD1+CTLA4+) cells among SARS-CoV-2-specific CD8+ T cells in severe relative to mild COVID-19 validated by manual gating. * p < 0.05 as determined by one-way analysis of variance with a Bonferroni post-test.

Examination of SARS-CoV-2-specific T cell clusters revealed one CD4+ and one CD8+ T cell cluster enriched in the severe group, and one CD4+ T cell cluster enriched in the mild group (Fig. 3G, 4G). The two clusters enriched in the severe group shared phenotypic features, including increased expression levels of the activation markers HLADR and CD38 and exhaustion markers PD1 and CTLA4, and decreased expression of the IL7 receptor alpha chain CD127 (Fig. 3H, 4H). These markers were not differentially expressed among the CD4+ cluster enriched in the mild group (Fig. 3H). To determine whether a subset of the markers identified by FlowSOM are sufficient to enrich for cells from the severe group, we manually gated on cells expressing the two activation/exhaustion markers PD1 and CTLA4. PD1+CTLA4+ cells were more frequent in the severe groups among both SARS-CoV-2-specific CD4+ and CD8+ T cells (Fig. 3I, 4I). These findings, together with the earlier observations based solely on manual gating, point to an overabundance of exhausted T cells recognizing SARS-CoV-2 spike during severe COVID-19.

### Recovery from severe COVID-19 associates with enhanced SARS-CoV-2-specific effector T cell response

Of our specimens analyzed by CyTOF, 33 were from SARS-CoV-2-infected individuals in the ICU, 32% of whom subsequently died from COVID-19. We next focused on these specimens to try to identify T cell signatures associated with survival from severe disease. Age did not account for the differential survival outcomes, as the ages of survivors and non-survivors were not significantly different (p = 0.8131) and the magnitude of the T cell response did not associate with age (R^2^ = 0.0032 among severe patients). Although male sex is a risk factor for severe COVID-19, we in fact found among our ICU patients a lower proportion of male patients that died (22%, compared to 40% for females). Furthermore, we found similar frequencies of SARS-CoV-2-specific T cells among our hospitalized male and female patients, and no marked phenotypic differences although there was a trend for higher activation of SARS-CoV-2-specific T cells in females (Fig. S4), consistent with previous reports of total HLADR+CD38+ T cells being higher in female COVID-19 patients (Takahashi et al., 2020). Because prior studies found high levels of SARS-CoV-2 antibodies to be associated with more severe disease (Garcia-Beltran et al., 2020; Liu et al., 2019; Woodruff et al., 2020), we assessed whether IgG and IgM against the SARS-CoV-2 spike and nucleocapsid were enriched in non-survivors. We did not find this to be the case, although we observed moderate/severe cases to harbor higher levels of antibodies than mild cases (Fig. S5).

We then examined whether the T cells from survivors and non-survivors differed in their response to spike peptide stimulation. Non-survivors more frequently failed to mount a robust SARS-CoV-2-specific T cell response, particularly for CD4+ T cells (Fig. 5A). The SARS-CoV-2-specific CD8+ T cell response trended higher in the survivors, but this did not reach statistical significance because of lower overall responses. Patients that mounted detectable SARS-CoV-2-specific T cell responses harbored significantly lower frequencies of total HLADR+CD69+ CD4+ and CD8+ T cells (Fig. S6), suggesting that an overall heightened state of T cell activation may hinder the development of T responses directed specifically against SARS-CoV-2.

**Figure 5.**
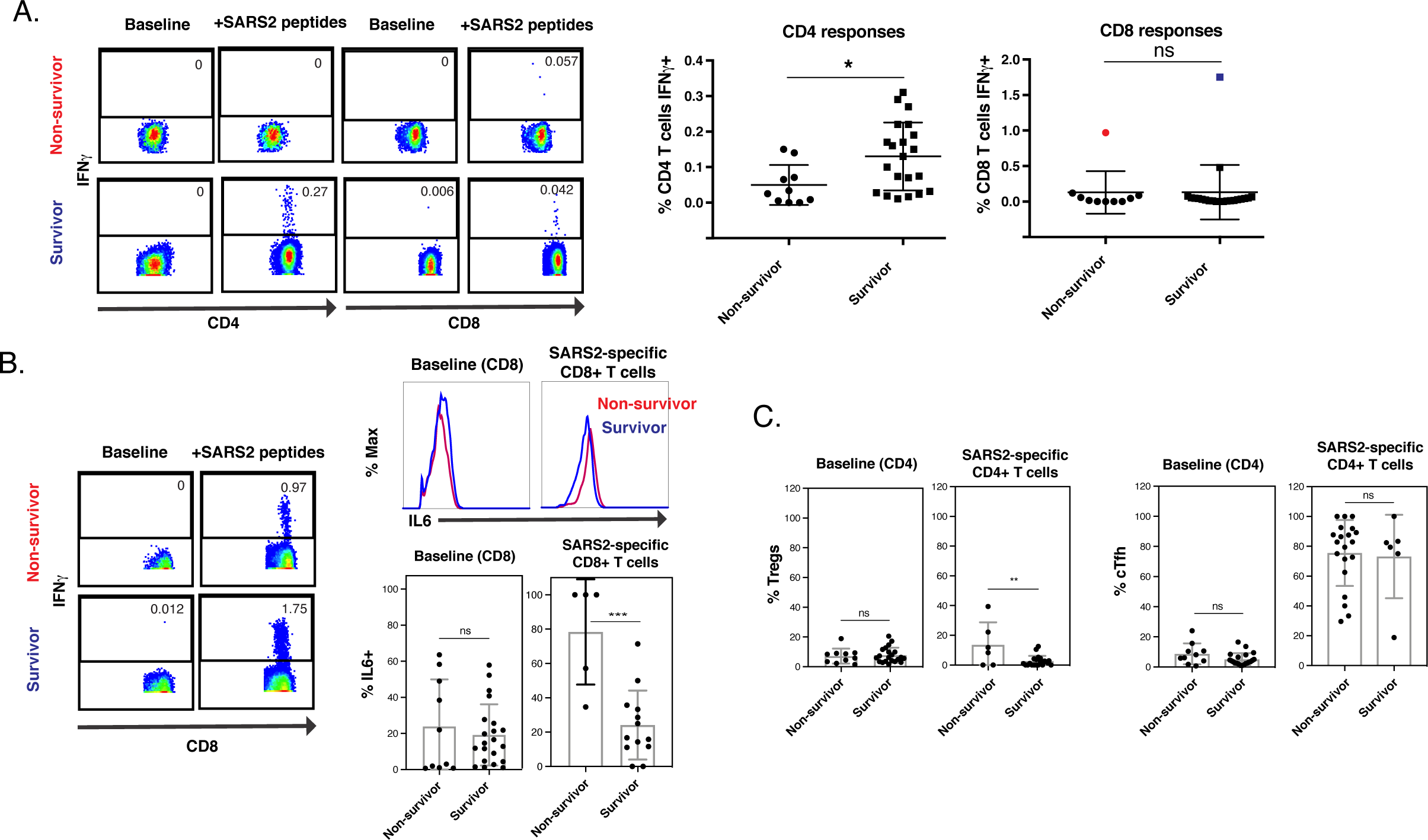
Distinct features of SARS-CoV-2-specific T cells associate with recovery from severe COVID-19. (**A**) Higher frequencies of SARS-CoV-2-specific T cells in individuals with severe COVID-19 that survived (“survivor”) than in those that did not (“non-survivor”). *Left:* Representative pseudocolor plots reflecting the percentage of SARS-CoV-2-specific CD4+ or CD8+ T cells from representative infected individuals in the ICU that did or did not survive COVID-19. *Right:* Quantification of the data shown in *(A)* across all infected ICU individuals. * p < 0.05, ns: non-significant as determined by a Student’s unpaired t-test. (**B**) SARS-CoV-2-specific CD8+ T cells from non-survivors produce elevated levels of IL6. *Left:* Pseudocolor plots of baseline and spike-stimulated samples from the two patients harboring the highest levels of SARS-CoV-2-specific CD8+ T cells in *panel A*, only one of whom survived. *Top right:* Histogram plots showing expression levels of IL6 in non-survivor and survivor CD8+ T cells. *Bottom right:* Proportion of IL6-producing cells among total and SARS-CoV-2-specific T cells in all non-survivor and survivor patients. (**C**) SARS-CoV-2-specific Tregs but not cTfh cells are significantly more abundant in non-survivors of severe COVID-19. See also Fig. S4 – S8.

Interestingly, two individuals mounted very robust CD8+ T cell responses to spike peptides, and one of these survived while the other did not (highlighted in blue and red, respectively, in Fig. 5A). After comparing all the antigens on these two patients’ CD8+ T cells, we found that IL6 levels within the SARS-CoV-2-specific CD8+ T cells were lower in the survivor than in the non-survivor, and this phenomenon was not observed among total CD8+ T cells (Fig. 5B). Of note, the subtleness of the increase in IL6 in SARS-CoV-2-specific CD8+ T cells from the non-survivor was not due to weak antibody staining, as a positive control of LPS-stimulated monocytes led to robust detection of IL6 (Fig. S7). This observation prompted us to examine IL6 expression among all the ICU individuals that did elicit SARS-CoV-2-specific CD8+ T cell responses. Indeed, we found a highly significant increase in IL6-producing SARS-CoV-2-specific CD8+ T cells in non-survivors, but not among total CD8+ T cells (Fig. 5B). These results suggest the possibility that SARS-CoV-2-specific CD8+ T cells producing both IFNγ and IL6 may contribute to lethal immunopathogenesis, although follow-up studies will be needed to confirm this hypothesis.

We next assessed whether the major T cell subsets were differentially distributed among the survivors and non-survivors. Memory CD4+ T cells were statistically more frequent in the survivors, but no statistically significant differences were observed among the Tcm, Tem, Ttm, Tn, Temra, or Tscm subsets (Fig. S8). Interestingly, however, SARS-CoV-2-specific Tregs were significantly more frequent among non-survivors, though total Tregs were not (Fig. 5C). In contrast, Tfh frequencies were equivalent between survivors and non-survivors (Fig. 5C).

Together, these observations are consistent with a model whereby Tregs recognizing SARS-CoV-2 spike can hinder the elicitation of a robust SARS-CoV-2-specific T cell response, and thus prevent recovery from severe disease.

### Clustering reveals more activated, long-lived SARS-CoV-2-specific CD4+ T cells in survivors of severe COVID-19

To compare T cells from survivors and non-survivors in an unbiased fashion, we subjected them to clustering via FlowSOM, and found phenotypic differences between the two groups for both CD4+ (Fig. 6A, B) and CD8+ (Fig. S9A, B) T cells. Among total CD4+ T cells, two clusters (B4.1 and B4.9) were enriched in the survivor group, although the former did not meet statistical significance (Fig. 6C). Both clusters consisted of activated (HLADR+) memory (CD45RO+) cells. While B4.1 expressed the additional activation markers CD38, CD69, and CCR5, B4.9 did not, consistent with earlier observations of HLADR being a more universal marker of T cell activation. Similar analyses among total CD8+ T cells revealed cluster B8.6 to be more frequent among survivors, albeit insignificantly after multiple correction (Fig. S9C). This cluster also harbored activated cells, and were predominantly Tem cells as suggested by low expression of CCR7 and CD62L. Interestingly, of all the CD8+ T cell clusters, this one harbored the highest levels of HLADR (Fig. S9D). These results support the concept that global CD4+ and CD8+ T cell activation, as assessed by canonical activation markers such as HLADR, is not detrimental but in fact, at least in our cohort, associated with survival from severe COVID-19.

**Figure 6.**
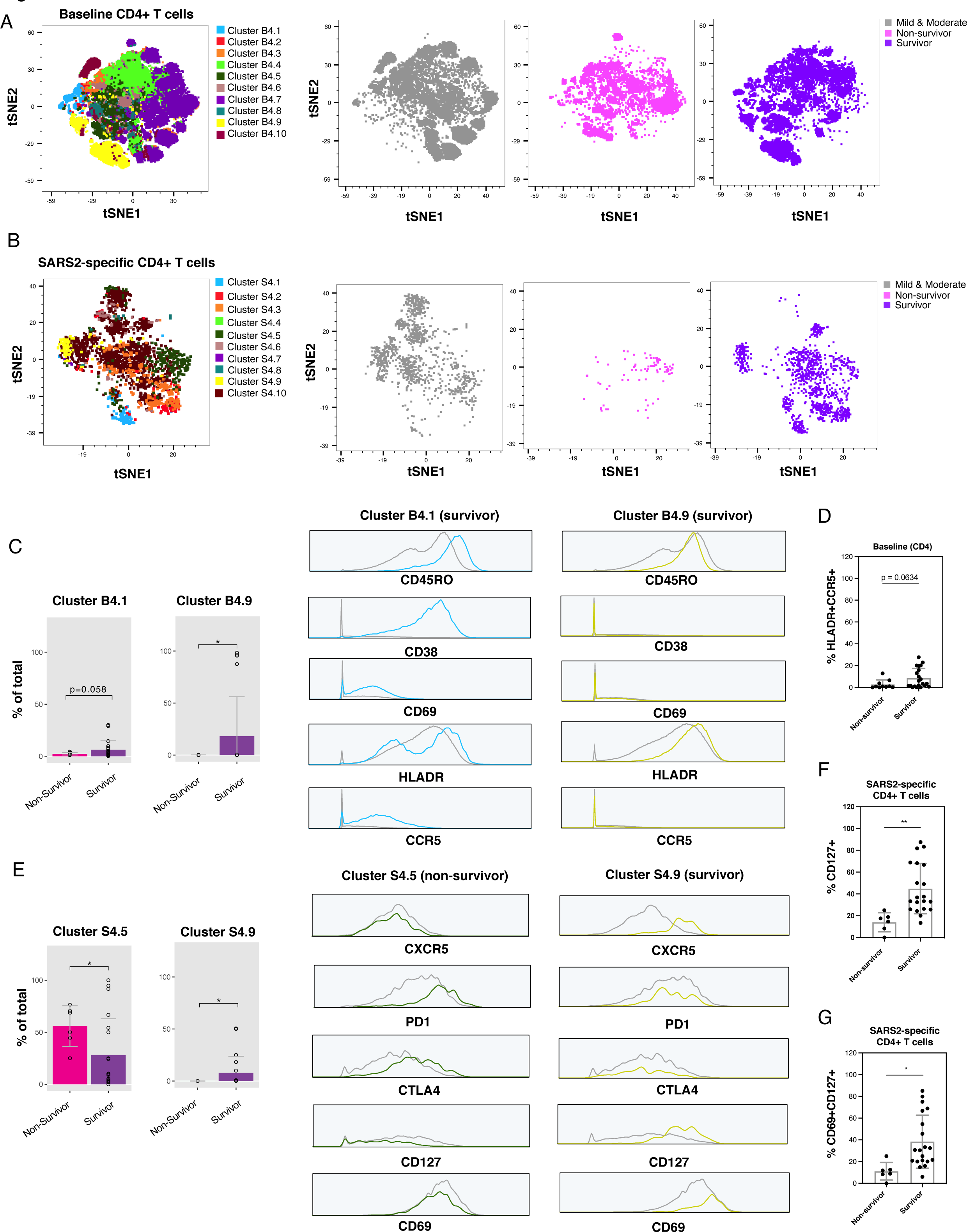
Clustering reveals enrichment of long-lived, activated SARS-CoV-2-specific CD4+ T cells in patients that successfully recover from severe COVID-19. (**A, B**) The phenotypes of total *(A)* and SARS-CoV-2-specific *(B)* CD4+ T cells differ between non-survivors and survivors of severe COVID-19. *Left:* tSNE plot and FlowSOM clusters identified in Fig. 3A *(A)* and Fig. 3C *(B)*. *Right:* Distribution of CD4+ T cells from non-survivors (*pink*), survivors (*purple*), and mild and moderate patient groups (*grey*). (**C**) Activated memory CD4+ T cells are elevated in survivors of severe COVID-19. *Left:* Clusters B4.1 and B4.9 are enriched in the survivor relative to the non-survivor group. * p < 0.05 as determined by a Student’s unpaired t-test. *Right:* Cells from clusters B4.1 and B4.9 are memory (CD45RO+) cells and express elevated levels of activation marker HLADR. Other activation markers (CD38, CD69, CCR5) are preferentially elevated only in cluster B4.1. Expression levels are displayed in grey for total CD4+ T cells and in color for clusters. (**D**) Survivors tend to harbor more activated (HLADR+CCR5+) CD4+ T cells. (**E**) *Left:* Cluster S4.5 is enriched in non-survivors, and cluster S4.9 in survivors. *Right:* Cells from cluster S4.5 co-express exhaustion markers PD1 and CTLA4, while cells from cluster S4.9 express high levels of cTfh marker CXCR5, low levels of exhaustion markers PD1 and CTLA4, high levels of IL7 receptor alpha chain CD127, and high levels of activation marker CD69. Expression levels from each cluster (*colored traces*) are shown against expression levels from all SARS-CoV-2-specific CD4+ T cell clusters combined (*grey traces*). * p < 0.05 as determined by a Student’s unpaired t-test. (**F, G**) SARS-CoV-2-specific CD4+ T cells expressing the homeostatic proliferation marker CD127 *(F)* or co-expressing CD127 with the activation marker CD69 *(G)* are more abundant in survivors than in non-survivors. * p < 0.05, ** p < 0.01, as determined by a Student’s unpaired t-test. See also Fig. S9 and S10.

This was further supported by our observation that activated CD4+ and CD8+ T cells identified by manual gating were significantly higher in the survivor than the non-survivor groups (Fig. 6D, S9E).

We then assessed the SARS-CoV-2-specific T cells. Among the CD4+ T cells, S4.5 was increased in non-survivors, while S4.9 was increased in the survivors. While S4.5 exhibited features of exhausted T cells (expressing high levels of PD1 and CTLA4), S4.9 exhibited features of cTfh cells with high proliferative potential (expressing high levels of CXCR5 and CD127) (Fig. 6E). Manual gating confirmed a significant increase in CD127-expressing SARS-CoV-2-specific CD4+ T cells in patients that survived severe disease (Fig. 6F). As cluster S4.9 also expressed high levels of activation marker CD69 (Fig. 6E), we assessed whether activated CD127+ cells were increased in survivors, and found this to indeed be the case (Fig. 6G).

Further analysis of cluster S4.9 confirmed it to be highly activated (expressing activation markers HLADR, CD38, CCR5, and Ox40 in addition to CD69), exhibit features of Tcm (expressing high levels of CCR7 and CD62L), and have mucosal tissue-homing potential (expressing high levels of CCR6 and CD49d) (Fig. S10). By contrast, cluster S4.5 did not harbor most of these features (Fig. S10). A cluster of SARS-CoV-2-specific CD8+ T cells significantly enriched in survivors was also identified (S8.2), and this cluster harbored phenotypic features distinct from the cluster of survivor-associated SARS-CoV-2-specific CD4+ T cells (Fig. S9F).

S8.2 was not activated (expressing low levels of HLADR and CD38) and, unlike S4.9, expressed low levels of CD127. Accordingly, unlike what was observed among SARS-CoV-2-specific CD4+ T cells, the percentages of CD127-expressing cells among SARS-CoV-2-specific CD8+ T cells were not increased among survivors (Fig. S9G). This suggests that relative to their CD4+ counterparts, SARS-CoV-2-specific CD8+ T cells in survivors may be less long-lived, although the observation that these cells express low levels of the terminal differentiation marker CD57 (Fig. S9F) suggests that they may be able to differentiate through non-homeostatic proliferative mechanisms.

### Escalating levels of SARS-CoV-2-specific T cells and dampening of bystander lung-homing activated cells predicts recovery from severe COVID-19

Finally, to better understand T cell features associated with survival of severe COVID-19, we assessed longitudinal specimens from our ICU cohort (Table S1). While survivors tended to increase the frequency of SARS-CoV-2-specific T cells over time, non-survivors tended to not have much of these cells to begin with, or the frequency of these cells stagnated or decreased over time (Fig. 7A, B). We then analyzed the high-dimensional phenotyping datasets of the longitudinal specimens for predictors of survival. Interestingly, total CD4+ and CD8+ T cell co-expressing CD69 and CXCR4 decreased over time in survivors, while the opposite was observed for non-survivors. Correspondingly, CD69-CXCR4-T cell frequencies increased in the days leading up to discharge from hospital, while they decreased in the days leading up to death (Fig. 7C). As CD69 is an activation marker and CXCR4 directs cells to various tissues, including the inflamed and damaged lung (Mamazhakypov et al., 2019), these observations support a model whereby activated bystander T cells recruited to the lung lead to poor disease outcome.

**Figure 7.**
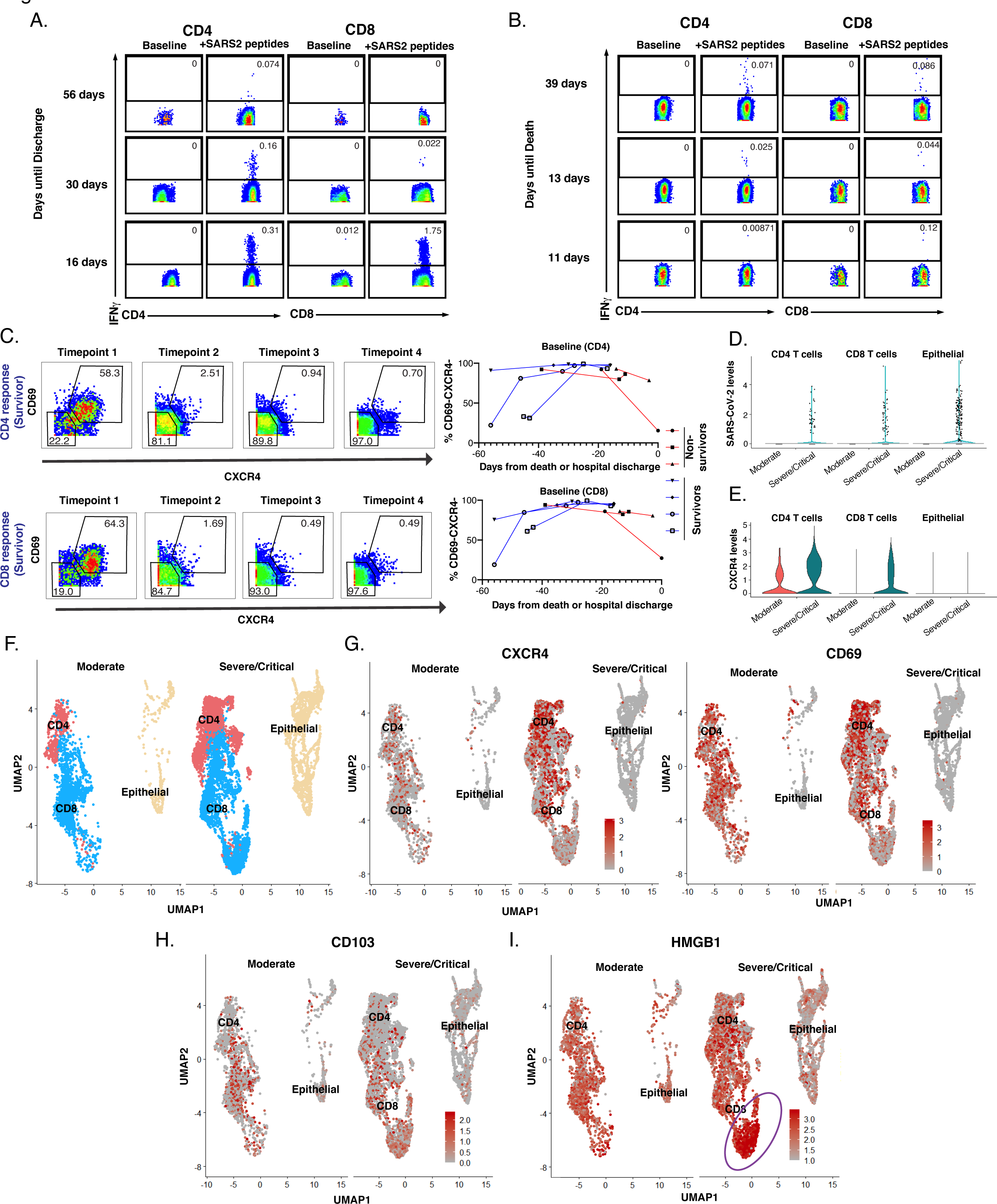
Escalating numbers of SARS-CoV-2-specific and diminishing numbers of lung-homing bystander CXCR4+ T cells correlate with survival of severe COVID-19. (**A, B**) Longitudinal analysis of a patient that survived *(A)* and did not survive (*B)* severe COVID-19, demonstrating an increasing number of SARS-CoV-2 T cells producing IFNγ over time in the survivor but not non-survivor. (**C**) Activated CXCR4-expressing T cells decrease over time in survivors but not non-survivors. *Left:* Pseudocolor plots showing gating for CD69+CXCR4+ and CD69-CXCR4-cells among total CD4+ and CD8+ T cells in longitudinal specimens from a representative survivor of severe COVID-19. *Right:* Longitudinal analyses of 7 cases of severe COVID-19, 4 of which survived infection. While the frequencies of CXCR4-CD69-T cells increase over time in survivors (*blue lines*), they decrease over time in non-survivors (*red lines*). Not shown are the opposite trend for CXCR4+CD69+ cells which over time decrease in survivors and increase in non-survivors. (**D**-**I**) Higher levels of infiltrating CXCR4+CD69+ T cells are present in lungs of severe/critical vs. moderate COVID-19 patients as determined by mining of public BAL scRNAseq datasets. Higher levels of SARS-CoV-2 viral reads in T and epithelial cells were detected during severe COVID-19 (*D*), which were also associated with higher levels of CXCR4 in T cells as depicted by violin plots (*E*). Note viral reads detected in T cells likely reflect cell-surface virion sticking as these cells do not express ACE2. UMAP visualization of T and epithelial cells from the scRNAseq datasets (*F*) revealed elevated expression of CXCR4 and CD69 on subsets of T cells (*G*) separate from T cells expressing high levels of the Trm marker CD103 (*H*). The CXCR4 ligand HMGB1 was expressed in T and epithelial cells, but especially concentrated among a subset of CXCR4-CD69-Trm CD8+ T cells outlined in purple (*I*). See also Fig. S11.

To provide further support for this model, we mined a public single-cell RNAseq (scRNAseq) dataset (Liao et al., 2020) of bronchoalveolar lavage (BAL) specimens from severe/critical COVID-19 patients, and compared it to BAL from moderate COVID-19 which harbored lower lung viral loads (Fig. 7D). Elevated expression of CXCR4 was observed in T but not epithelial cells of severe patients (Fig. 7E). Visualization of the T cells by UMAP (Fig. 7F, S11A) revealed co-expression of CXCR4 and CD69 in the same subsets (Fig. 7G). These cells likely infiltrated from the periphery as they express low levels of the T resident memory (Trm) marker CD103 (Fig. 7H). To assess whether pulmonary T cells may themselves recruit more CXCR4+ T cells, we assessed for expression of the CXCR4 ligands CXCL12 and HMGB1 (Schiraldi et al., 2012). While CXCL12 was not expressed (Fig. S11B), HMGB1 was, particularly in a subset of CD8+ T cells over-represented in severe patients (Fig. 7I). These CD8+ T cells comprise a unique cluster (Fig. S11C) and express relatively low levels of CXCR4 but high levels of CD103 suggesting they are mostly Trm. Together, these data suggest that during severe COVID-19, activated bystander T cells may, through expression of CXCR4, be recruited to the lung by HMGB1 produced by pulmonary cells particularly CD8+ Trm cells, which could contribute to COVID-19-associated mortality.

## DISCUSSION

T cell lymphopenia was identified early on during the COVID-19 pandemic as a hallmark of severe disease, implying an important role for T cells in the control of SARS-CoV-2. Yet, the T cell subsets that may contribute to recovery and the role of T cells directly recognizing SARS-CoV-2 epitopes has not been investigated in depth. In this study, we compared total and SARS-CoV-2-specific T cells from mild, moderate, and severe cases of COVID-19, and within the severe cases conducted in-depth analyses of longitudinal specimens to identify features predicting who will survive severe COVID-19. As discussed below, we discovered T cell features associated with recovery from disease, but also some implicated in disease pathogenesis. As part of our study, we provide as a resource the raw CyTOF single-cell datasets for all the total and SARS-CoV-2-specific T cells for further data mining by the research community (see methods).

### Elevated levels of activated and exhausted T cells during severe COVID-19

The phenotypes of total T cells from COVID-19 patients from our study are largely consistent with a recent study that conducted deep-profiling of immune cells (including T cells) using high-parameter flow cytometry (Mathew et al., 2020). We, like the prior study, had observed higher frequencies of CD8+ Ttm cells in mild patients, and a higher frequency of activated, PD1-expressing T cells in severe patients. We also, similar to multiple other studies (Mathew et al., 2020; Rydyznski Moderbacher et al., 2020), found memory T cells to be significantly more abundant in severe patients, although this was likely due to the older age of the severe group, which affects memory cell frequencies (Rydyznski Moderbacher et al., 2020).

Among SARS-CoV-2-specific cells, CD4+ Tem cells were increased in severe cases, but CD4+ Temra cells were markedly depleted. The biological significance of this difference is unclear, as both Tem and Temra cells can play important roles in antiviral immunity (Dunne et al., 2002; Lilleri et al., 2008; Northfield et al., 2007; Sridhar et al., 2013; Weiskopf et al., 2015).

SARS-CoV-2-specific cTfh cells were also more abundant in moderate and severe cases, and this paralleled a corresponding increase in total cTfh cells in more severe cases, the latter of which was previously reported (Oja et al., 2020).

T cell exhaustion as defined by the exhaustion marker PD1 was reported early during the pandemic (De Biasi et al., 2020; Diao et al., 2020) but a recent study suggested that PD1+

CD8+ T cells from convalescent COVID-19 patients can be functional and that PD1 may more often represent an activation marker in these individuals (Rha et al., 2020). We therefore looked within our datasets for features of T cell exhaustion among our total and SARS-CoV-2-specific cells. We defined exhausted cells as those dually expressing elevated PD1 and TIGIT, or PD1 and CTLA4. In both instances we found a significant increase in exhausted SARS-CoV-2-specific T cells in severe cases. These results, together with the finding that T cells from hospitalized patients activated by SARS-CoV-2 peptide stimulation express elevated transcript levels of multiple exhaustion markers (PD1, TIGIT, LAG3, TIM3) relative to those from non-hospitalized patients (Meckiff et al., 2020), suggest that T cell exhaustion correlates with disease severity. We also observed co-expression of PD1 with the Fas receptor CD95 involved in apoptosis, and that PD1+CD95+ T cells, within both the total and SARS-CoV-2-specific compartments, were elevated in severe relative to mild cases. These observations are in line with an upregulation of CD95 transcripts on total T cells during COVID-19 (Zhu et al., 2020), and with reports of apoptotic T cells during severe (Yao et al., 2020) and fatal (Feng et al., 2020) COVID-19, during which elevated CD95 was observed in the dying cells. Together these results suggest increased proportions of exhausted and apoptotic-prone T cells during severe COVID-19.

### SARS-CoV-2-specific T cell frequency predicts survival from severe COVID-19

To our knowledge, this is the first study to directly compare via in-depth immune phenotyping the features of T cells from severe (ICU) COVID-19 patients that do vs. do not survive disease. We discovered in a cross-sectional analysis that survivors mounted a higher SARS-CoV-2-specific T cell response, and longitudinal analyses revealed that this response increases in survivors prior to recovery, whereas it does not in non-survivors. These data suggest that SARS-CoV-2-specific T cells are protective during severe COVID-19, and are in line with a number of other reports, including: a recent report of greater expansion of SARS-CoV-2-specific T cells during moderate than severe COVID-19 (Liao et al., 2020); the finding that antigen-specific T cells against SARS-CoV-1, a close relative of SARS-CoV-2, are protective in mouse infection models (Zhao et al., 2016); and a recent study demonstrating SARS-CoV-2-specific T cell responses, as defined by AIM markers, to be associated with less severe disease (Rydyznski Moderbacher et al., 2020). Although that latter study did not focus on fatal vs. non-fatal outcomes, it included one fatal case with longitudinal sampling, and observed that patient to have had no detectable SARS-CoV-2-specific T cell responses 16 and 11 days prior to death, in line with our findings. Of note, however, a separate research group reported the opposite trend—AIM-identified SARS-CoV-2-specific T cells associating with more severe disease—although the results did not meet statistical significance (Anft et al., 2020; Thieme et al., 2020). Regardless, our data comparing fatal vs. non-fatal severe COVID-19 are consistent with the notion SARS-CoV-2-specific T cells being beneficial rather than detrimental for surviving severe COVID-19. That being said, we did find that SARS-CoV-2-specific CD8+ T cells co-producing IFNγ and IL6 were elevated in individuals that did not survive severe disease. Plasma IL6 levels associate with COVID-19 severity (Huang et al., 2020; Mathew et al., 2020; Zhou et al., 2020) and predict COVID-19-associated death (Del Valle et al., 2020). However, save a study that reported elevated IL6 production in total CD4+ T cells from COVID-19 patients in the ICU, IL6 is not thought to be produced by T cells during SARS-CoV-2 infection. Whether IL6 production by SARS-CoV-2-specific CD8+ T cells directly contributes to immunopathogenesis remains to be determined.

### Potential mechanisms underlying diminished SARS-CoV-2-specific T cell responses in non-survivors of severe COVID-19

Our data suggest multiple mechanisms by which severe patients may control the production of SARS-CoV-2-specific T cells. Hospitalized patients that produced few SARS-CoV-2-specific T cells—and were more likely to die—harbored higher frequencies of SARS-CoV-2-specific Tregs. Although little is known about SARS-CoV-2-specific Tregs, the proportion of Tregs among total T cells has been reported to be both increased (De Biasi et al., 2020) and decreased (Qin et al., 2020) during severe COVID-19. Consistent with increased levels of Tregs during severe infection are observations of disease severity positively associating with levels IL10, a regulatory cytokine commonly produced by Tregs (Han et al., 2020; Zhao et al., 2020b). We did not observe increased levels of total Tregs in fatal vs. non-fatal cases of severe COVID-19, but the increased number of SARS-CoV-2-specific Tregs in non-survivors may suffice to hinder the differentiation of SARS-CoV-2-specific effector T cells.

Our finding may seem at odds with a recent study suggesting a decrease in SARS-CoV-2-specific Tregs during severe disease (Meckiff et al., 2020). However, differences in experimental design likely account for the differences. While our study compared ICU patients that survived vs. did not survive, the other study compared ICU patients to non-hospitalized ones. Furthermore, the prior study identified SARS-CoV-2-specific Tregs as those expressing AIM markers after stimulating for 24 hours with SARS-CoV-2 peptides, while we identified the cells as those producing IFNγ after 6 hours of stimulation. As all the SARS-CoV-2-specific cells characterized in our study were defined based on IFNγ induction, the SARS-CoV-2-specific Tregs we identified are in fact a subset of all possible Tregs. These IFNγ+ Tregs have been reported to play important roles during graft rejection and autoimmunity (Daniel et al., 2008; McClymont et al., 2011), but to our knowledge have not been described during viral infections.

Another potential mechanism accounting for increased frequencies of SARS-CoV-2-specific T cells in survivors is their harboring increased expression of CD127, a component of the IL7 receptor complex important for homeostatic proliferation. We recently demonstrated that SARS-CoV-2-specific T cells from convalescent patients homeostatically proliferate in response to IL7 signaling (Neidleman et al., 2020a), demonstrating the expansion potential of these cells. We speculate that during severe COVID-19, elevated expression of CD127 allows SARS-CoV-2-specific T cells to expand more rapidly and thereby persist in sufficient quantities to aid in recovery.

### CXCR4 antagonism as target for T cell-mediated immunopathogenesis in the lung

One of the most interesting findings of this study came out of our analysis of longitudinal specimens from fatal vs. non-fatal severe cases, where we found CXCR4+CD69+ T cells decreasing over time in those that survive, and increasing in those that do not. CXCR4 plays important roles in lymphocyte chemotaxis and in homing of hematopoietic stem cells to bone marrow. Its primary ligand, CXCL12, is expressed in multiple tissues, but of importance for this study, is elevated in the damaged lung, particularly the lung vasculature (Mamazhakypov et al., 2019). Interestingly, CD69+CXCR4+ T cells have been found at elevated levels in non-small cell lung cancer tissues as compared to normal lung tissue (Wald et al., 2006), suggesting their increased presence under pathologic conditions. Indeed, we found that T cells expressing CXCR4 and CD69 are in fact elevated in BAL of severe COVID-19 patients. These cells likely migrated from the periphery as they express low levels of the Trm marker CD103. Consistent with an infiltration of pathogenic T cells during severe COVID-19 are observations that pulmonary T cells during critical as compared to moderate disease expressed fewer tissue-resident transcripts (Liao et al., 2020) and proteins (Oja et al., 2020). Trm cells may, however, play a role in pathogenesis by producing the CXCR4 ligand HMGB1, which, consistent with our findings, has been reported to be elevated during fatal COVID-19 (Chen et al., 2020b).

Together with the observation of activation of CMV- and EBV-specific T cells during the acute phase of COVID-19 (Sekine et al., 2020), these findings bring up the possibility that during critical and fatal COVID-19, T cells in the periphery activated through bystander effects are recruited to the lung via CXCR4-mediated chemotaxis, and this contributes to pulmonary damage. If CXCR4-driven T cell infiltration does indeed contribute to fatal COVID-19, then inhibitors against this receptor may be useful. AMD3100, a CXCR4 antagonist that has been used to mobilize stem cells (Cashen et al., 2007), could be tested in clinical trials. Of note, optimized derivatives of a peptide called EPI-X4, originally discovered by virtue of its ability to inhibit CXCR4-tropic HIV infection of target cells (Zirafi et al., 2015), exhibit superior activity over AMD3100 for blocking CXCR4-mediated chemotaxis, and have been shown to potently block migration of immune cells into the lungs of asthmatic mice (Harms et al., 2020). These derivatives represent intriguing candidates for treatment of severe COVID-19.

### Conclusions

In summary, our findings overall support a beneficial rather than immunopathologic role for effector SARS-CoV-2-specific T cells during severe acute infection. This was evidenced by survivors of severe infection exhibiting: 1) escalating SARS-CoV-2-specific T cell numbers before recovery, 2) increased levels of SARS-CoV-2-specific T cells with homeostatic proliferative potential, and 3) diminished numbers of immunosuppressive SARS-CoV-2-specific Tregs. The mechanisms by which T cells would help recovery from severe infection is not clear, but likely involves providing B cell help and direct cytolysis of infected cells. In addition, as IFNγ downregulates ACE2 (de Lang et al., 2006), production of this cytokine by SARS-CoV-2-specific T cells may limit the availability of host cells for the virus. Exceptions to the notion of “beneficial” T cells, however, are our observations of non-survivors expressing elevated levels of IL6-producing SARS-CoV-2-specific CD8+ T cells, and the increase of activated, lung-homing bystander T cells in the days leading up to death. Nevertheless, taking into consideration the longevity of SARS-CoV-2-specific T cells (Zuo et al., 2020), our results suggest that strategies to boost the effector functions of SARS-CoV-2-specific T cells, including by vaccination, will be beneficial for decreasing COVID-19 mortality and helping to end this devastating pandemic.

### Limitations

Our study has limitations. To enable deep-phenotyping of SARS-CoV-2-specific T cells, we had to characterize a relatively small cohort of 34 infected individuals. This may have accounted for our inability to detect sex-based differences. A second limitation was our use of peptide stimulation to identify SARS-CoV-2-specific cells. The phenotypes of our SARS-CoV-2-specific T cells therefore correspond to those immediately following antigen encounter, rather than at baseline. Importantly, however, we did not compare the phenotypes of baseline cells to SARS-CoV-2-specific ones, as such a comparison would include artifacts resulting from the stimulation. Instead, we always compared SARS-CoV-2-specific T cells between different patient groups. Future studies using tetramers will be needed to assess the phenotypes of baseline SARS-CoV-2-specific T cells along the entire spectrum of COVID-19 disease. Such studies, however, would be limited to analyzing responses against a small number of epitopes, and would be for the most part limited to CD8+ T cells as tetramer reagents for CD4+ T cells are generally not reliable. A third limitation was our restricting analysis of SARS-CoV-2-specific T cells to those recognizing the spike protein, which elicits a limited CD8+ T cell response. Future studies should examine responses against proteins more commonly recognized by SARS-CoV-2-specific CD8+ T cells (Ferretti et al., 2020). Fourthly, this study was a phenotyping and correlative study, so follow-up functional assays will be required to establish cause vs. effect.

Finally, our CyTOF analysis was limited to cells from the blood, although we confirmed the presence of activated CXCR4+ T cells in the lungs of severe COVID-19 patients.

## Data Availability

Datasets are available through the following link: https://doi.org/10.7272/Q67H1GTB

## ACKNOWLEDGEMENTS

This work was supported by the Van Auken Private Foundation and David Henke (N.R.R.); the Program for Breakthrough Biomedical Research (N.R.R., E.G., S.L., J.V., which is partly funded by the Sandler Foundation; philanthropic funds donated to Gladstone Institutes by The Roddenberry Foundation and individual donors devoted to COVID-19 research (N.R.R.); Awards #2164 (N.R.R.), #2208 (N.R.R.), and #2160 (to S.L.) from Fast Grants, a part of Emergent Ventures at the Mercatus Center, George Mason University; and NIH R01 AI123126-05S1 (E.G.). We acknowledge the NIH DRC Center Grant P30 DK063720 and the S10 1S10OD018040-01 for use of the CyTOF instrument. We also thank Stanley Tamaki, Tomoko Kakegawa Peech, and Claudia Bispo for CyTOF assistance at the Parnassus Flow Core, Heather Hartig for help with recruitment, Nicole Lazarus and Eugene Butcher for the Act1 antibody, John C.W. Carroll for graphics arts, Françoise Chanut for editorial assistance, and Robin Givens for administrative assistance.

## AUTHOR CONTRIBUTIONS

J.N., and A.G. processed specimens, designed and performed experiments, and conducted data analyses; X.L. and J.Y. conducted data analyses; M.M. processed samples and conducted data analyses; C.Y. collected in-patient specimens, conducted patient chart analyses, and performed experiments; V.M. and G.G. conducted CHIRP participant interviews, enrollment, and specimen collection; W.C.G. participated in data analysis and performed supervision; J.V. conceived ideas for the study; E.G. conceived ideas for the study, performed supervision, and conducted data analyses; S.L. established the CHIRP cohort and conducted CHIRP participant interviews, enrollment, and specimen collection; K.L. performed supervision, conducted data analyses, and provided specimens; N.R.R. conceived ideas for the study, performed supervision, conducted data analyses, and wrote the manuscript. All authors read and approved the manuscript.

## COMPETING FINANCIAL INTERESTS

The authors declare no competing financial interests.

## METHODS

### RESOURCE AVAILABILITY

#### Lead Contact

Requests for resources and reagents and for further information should be directed to and will be fulfilled by the Lead Contact, Nadia Roan.

#### Materials Availability

This study did not generate new unique reagents.

#### Data and Code Availability

The raw CyTOF datasets generated from this study are available for download through the public repository Dryad via the following link: https://doi.org/10.7272/Q67H1GTB

### EXPERIMENTAL MODEL AND SUBJECT DETAILS

#### Human Subjects

Blood was drawn from 34 SARS-CoV-2-infected and 11 uninfected individuals. Infection status was established by RT-PCR. Of the infected individuals, 19 experienced a severe course of disease as defined by being in the ICU (designated “survivor” in this study), and 6 of these individuals died from COVID-19 (designated “non-survivor” in this study). Moderate cases were defined as non-ICU hospitalizations for COVID-19. Eight of the individuals that experienced severe disease were sampled longitudinally at up to 4 timepoints. All specimens from hospitalized patients were from a timepoint when the patients were still hospitalized, and this ranged from 0-76 days after initial positive SARS-CoV-2 test. Mild cases consisted of those never hospitalized for COVID-19, and were generally analyzed 20-154 after initial positive SARS-CoV-2 test. While specimens from mild, moderate, and severe cases may have been collected at similar times after infection, the specimens from the moderate and severe cases were from hospitalized individuals, and are therefore considered acute specimens. By contrast, specimens from the mild cases were from outpatient individuals that were no longer symptomatic at the time of sampling and therefore can be considered convalescent specimens. Additional clinical features, including patient gender, age, race, and whether patients were given convalescent plasma, dexamethasone, or remdesivir for COVID-19, are indicated in Table S1. Consistent with male sex, advanced age, and non-white race being risk factors for severe disease, mild cases were predominantly female (81%, vs. 48% for hospitalized patients), younger (median age 41, vs. 64 for hospitalized patients), and all of white ethnicity (vs. 72% LatinX for hospitalized patients). The demographics of severe fatal vs. non-fatal cases of COVID-19 were more closely matched. In our cohort, fatal cases were more frequently female (67%, vs. 54% for non-fatal), older (median age 73, vs. 64 for non-fatal), and LatinX (100%, vs. 61.5% for non-fatal). Hospitalized patients were all from the Zuckerberg San Francisco General Hospital (ZSFG), while out-patients were recruited from the COVID-19 Host Immune Pathogenesis (CHIRP) study. This study was approved by the University of California, San Francisco (IRB # 20-30588).

### METHOD DETAILS

#### Preparation of specimens for CyTOF

On the day of each blood draw, PBMCs were isolated from blood using Lymphoprep^TM^ (StemCell Technologies). Six million cells were then immediately treated with cisplatin (Sigma-Aldrich) as a live/dead marker and fixed with paraformaldehyde (PFA) as previously described (Ma et al., 2020; Neidleman et al., 2020a), unless there were insufficient cell numbers in which case fewer cells were treated in this manner. Fixation was carried out by resuspending cells in 2 ml PBS (Rockland) with 2 mM EDTA (Corning), and then adding 2 ml of PBS containing 2 mM EDTA and 25 μM cisplatin (Sigma-Aldrich). After 1 min of incubation, cisplatin staining was quenched with 10 ml of CyFACS (metal contaminant-free PBS (Rockland) supplemented with 0.1% FBS and 0.1% sodium azide (Sigma-Aldrich)). The cells were then centrifuged, resuspended in 2% PFA in CyFACS, and incubated for 10 minutes at room temperature. After two more washes with CyFACS, cells were resuspended in CyFACS containing 10% DMSO and stored at -80°C until analysis by CyTOF. These cells constituted the “baseline” specimens.

The remaining cells were stimulated for 6 hours with the co-stimulatory agents 0.5 μg/ml anti-CD49d clone L25 and 0.5 μg/ml anti-CD28 clone L293 (both from BD Biosciences) and 0.5 μM of overlapping 15-mer SARS-CoV-2 spike peptides PepMix™ SARS-CoV-2 Peptide (Spike Glycoprotein) (JPT Peptide Technologies) in RP10 media (RPMI 1640 medium (Corning) supplemented with 10% fetal bovine serum (FBS, VWR), 1% penicillin (Gibco), and 1% streptomycin (Gibco)). To test the functionality of the IL6 antibody, PBMCs were stimulated instead with 200 ng/ml lipopolysaccharide LPS (Sigma-Aldrich). A final concentration of 3 μg/ml Brefeldin A Solution (eBioscience) was also included to enable detection of intracellular cytokines. For samples that had sufficient cell yield, a positive control of stimulation with 16 nM PMA (Sigma-Aldrich) and 1 μm ionomycin (Sigma-Aldrich) was additionally included. Treated cells were then cisplatin-treated and PFA-fixed as described above.

#### CyTOF staining and data acquisition

CyTOF staining was conducted using recently described methods (Cavrois et al., 2017; Ma et al., 2020; Neidleman et al., 2020a; Neidleman et al., 2020b). Cisplatin-treated cells were thawed and washed with CyFACS in Nunc 96 DeepWell^TM^ polystyrene plates (Thermo Fisher) at a concentration of 6 x 10^6^ cells / 800 μl in each well. Cells were then blocked with mouse (Thermo Fisher), rat (Thermo Fisher), and human AB (Sigma-Aldrich) sera for 15 minutes at 4°C. Samples were then washed twice in CyFACS and stained at 4°C for 45 minutes with surface CyTOF antibodies (Table S2) in a final volume of 100 μl. The samples were then washed three times with CyFACS, and fixed overnight at 4°C in 100 μl of 2% PFA in PBS. Samples were then washed twice with Intracellular Fixation & Permeabilization Buffer (eBioscience) and incubated for 45 minutes at 4°C. After two additional washes with Permeabilization Buffer (eBioscience), samples were blocked for 15 minutes at 4°C in 100 μl of mouse and rat sera diluted in Permeabilization Buffer. After another round of washing with Permeabilization Buffer, samples were stained at 4°C for 45 minutes with intracellular CyTOF antibodies (Table S2) in a final volume of 100 μl. The cells were then washed with CyFACS, and stained for 20 minutes at room temperature with 250 nM of Cell-ID^TM^ Intercalator-IR (Fluidigm). Immediately prior to sample acquisition, cells were washed twice with CyFACS buffer, once with MaxPar® cell staining buffer (Fluidigm), and once with Cell acquisition solution (CAS, Fluidigm), and then resuspended in EQ™ Four Element Calibration Beads (Fluidigm) diluted in CAS. Samples were acquired on a Helios-upgraded CyTOF2 instrument (Fluidigm) at the UCSF Parnassus flow core facility.

### QUANTIFICATION AND STATICAL ANALYSIS

#### CyTOF data analysis

CyTOF datasets were exported as flow cytometry standard (FCS) files, and de-barcoded according to manufacturer’s instructions (Fluidigm). FlowJo software (BD Biosciences) was used to identify CD4+ T cells (live, singlet CD3+CD19-CD4+CD8-) and CD8+ T cells (live, singlet CD3+CD19-CD4-CD8+) among the baseline and stimulated samples. Peptide-stimulated samples were further separated into bystander (IFNγ-) and SARS-CoV-2-specific (IFNγ+) T cells. For subset and high-dimensional analyses of SARS-CoV-2-specific T cells, we excluded patients with fewer than three SARS-CoV-2-specific T cells to limit skewing of data. All subset percentages were reported relative to number of live, singlet PBMCs, or the indicated T cell subset. t-SNE analyses were performed using the Cytobank software with default settings. All phenotyping cellular markers not used in the upstream gating strategy were included in generating the t-SNE plots. Non-cellular markers (e.g., live/dead stain) and cytokines were excluded for the generation of t-SNE plots. Dot plots were generated using both Cytobank and FlowJo. Clustering was conducted using the unsupervised clustering algorithm FlowSOM (Van Gassen et al., 2015) within Cytobank, using default settings (Clustering method: Hierarchical consensus; Iterations: 10; Seed: Automatic). The same parameters used in t-SNE plot generation were used as FlowSOM clustering parameters. The statistical tests used in comparison of groups are indicated within the figure legends.

#### Antibody Measurement

IgM and IgG antibodies against SARS-CoV-2 spike receptor binding domain (RBD) and nucleocapsid protein (NP) were measured in samples from patients that did not receive convalescent plasma, using the Pylon 3D automated immunoassay system (ET Healthcare) as described (Lynch et al., 2020). Briefly, quartz glass probes precoated with either affinity-purified goat anti-human IgM (to capture IgM) or Protein G (to capture IgG) were dipped into 15 μl of diluted patient plasma. After washing, the probe was dipped into the assay reagent containing both biotinylated recombinant RBD and NP. After additional washes, the probe was incubated with a Cy5-conjugated streptavidin polysaccharide conjugate reagent. The background-corrected signal was reported as relative fluorescent units (RFU), reflecting the relative levels of specific antibodies in each specimen.

#### Analysis of public scRNAseq datasets of COVID-19 bronchoalveolar lavage specimens

FASTQ files generated from BAL samples of two moderate (GSM4339769, GSM4339770) and six severe/critical patients were downloaded from the Gene Expression Omnibus (GEO) database under accession code GSE145926 (https://www.ncbi.nlm.nih.gov/geo/query/acc.cgi?acc=GSE145926) (Liao et al., 2020). These scRNAseq datasets were generated using the Chromium Single Cell 5’ library preparation kit (10X Genomics). Cell Ranger software version 3.1.0 (10x Genomics) was used to generate the filtered cell-barcode matrices. A standard Seurat v.3 workflow (Stuart et al., 2019) was used to generate cell clusters after filtering out dead cells (those containing more than 10% of mitochondrial gene counts). T and epithelial cell clusters were then extracted from each sample, by identification of clusters expressing the CD3 and EPACM genes, respectively. The filtered gene-barcode matrices for the extracted cells were normalized using ‘LogNormalize’ with default parameters. The top 2,000 variable genes were then identified using the ‘vst’ method by the ‘FindVariableFeatures’ function. To integrate the 8 BAL samples and remove potential batch effects, we used the standard integration Seurat v.3 workflow. The first 30 dimensions from canonical correlation analysis (CCA) were used as the input parameter for the ‘FindTransferAnchors’ function. Principal component analysis (PCA) was performed using the top 2,000 variable genes of the integrated matrix, followed by implementation of the graph-based clustering algorithm on the PCA-reduced data. Finally, UMAP visualizations were generated using the top 30 PCA. The UMAP resolution was set to 0.8. To detect SARS-CoV-2 transcripts, a customized reference genome/transcriptome was built by integrating the both human GRCh38 and SARS-CoV-2 genomes (severe acute respiratory syndrome coronavirus 2 isolate Wuhan-Hu-1, complete genome, GenBank MN908947.3 (https://www.ncbi.nlm.nih.gov/nuccore/MN908947.3/) and FASTQ alignment was performed using the splice-aware aligner STAR (Dobin et al., 2013).

## SUPPLEMENTARY FIGURE LEGENDS

**Figure S1.**
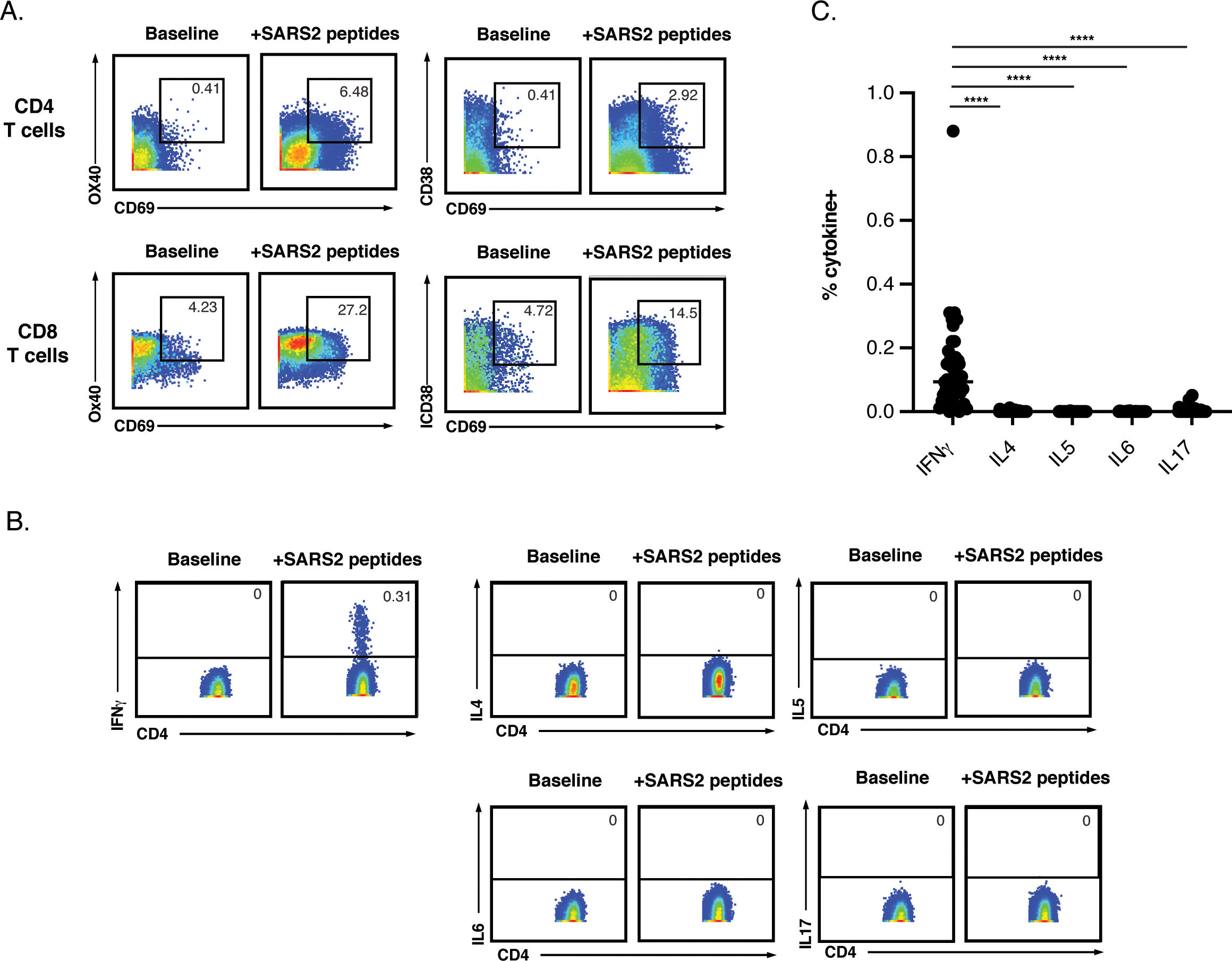
Assessment of SARS-CoV-2-specific T cells by AIM and cytokine production – Related to Figure 1. (**A-C**) SARS-CoV-2-specific T cells were assessed for the production of activation-induced markers (AIM) in pairs (*A*), or of individual cytokines IFNγ, IL4, IL5, and IL17 (*B, C*) in the absence or presence of stimulation with overlapping 15-mers spanning SARS-CoV-2 spike. (**A**) The AIM markers were assessed in baseline and SARS-CoV-2 peptide-stimulated samples from a representative severe COVID-19 patient. Although AIM markers were upregulated upon peptide stimulation, they were also expressed at substantial levels in the baseline samples, and responding cells could not be clearly distinguished from non-responding cells based on AIM expression. Numbers refer to the percentages of cells within the corresponding gates. (**B**) Peptide stimulation of CD4+ T cells from the donor from *panel A* induced the production of IFNγ, but not IL4, IL5, IL6, and IL17. Numbers refer to the percentages of cells within the corresponding gates. (**C**) Cumulative data from all donors showing the percentages of cytokine+ cells following stimulation with SARS-CoV-2 peptides. **** p < 0.0001 as determined by a one-way analysis of variance with a Bonferroni post-test.

**Figure S2.**
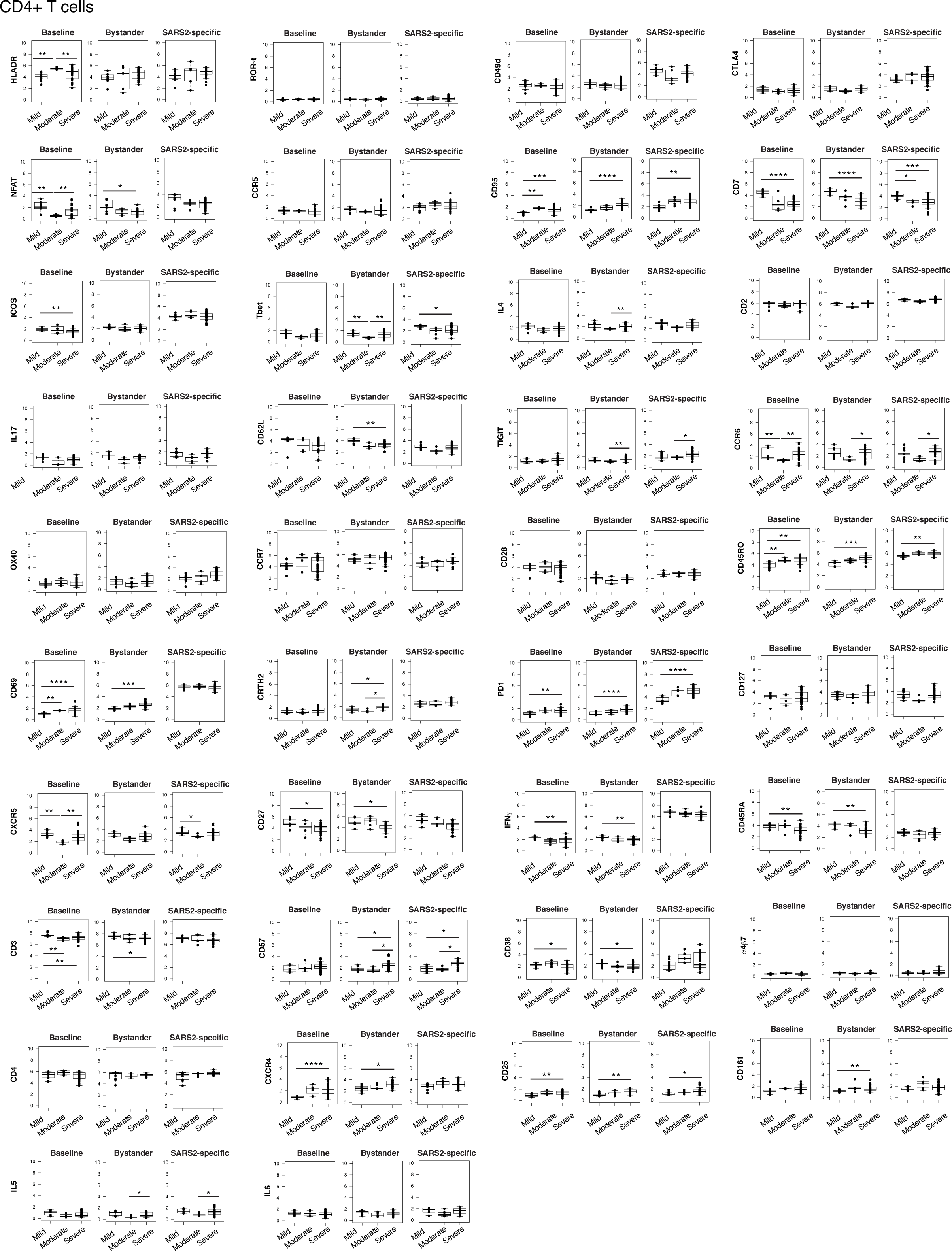
Mean signal intensity reflecting expression levels of 38 surface and intracellular antigens in baseline, bystander, and SARS-CoV-2-specific CD4+ T cells from mild, moderate, and severe COVID-19 – Related to Figure 1. PBMCs were purified from freshly drawn blood specimens and phenotyped by CyTOF at baseline or following a 6-hour stimulation with anti-CD49d/CD28 and overlapping 15-mer peptides from SARS-CoV-2 spike. Bystander cells are cells that remained IFNγ-in the stimulated samples. SARS-CoV-2-specific CD4+ T cells are the cells that produced IFNγ following stimulation. Shown are the mean signal intensity (MSI) values for arcsinh-transformed data. *p < 0.05, **p < 0.01, ***p < 0.001, ****p < 0.0001 as assessed using the Student’s unpaired t-test and adjusted for multiple testing using the Benjamini-Hochberg for FDR.

**Figure S3.**
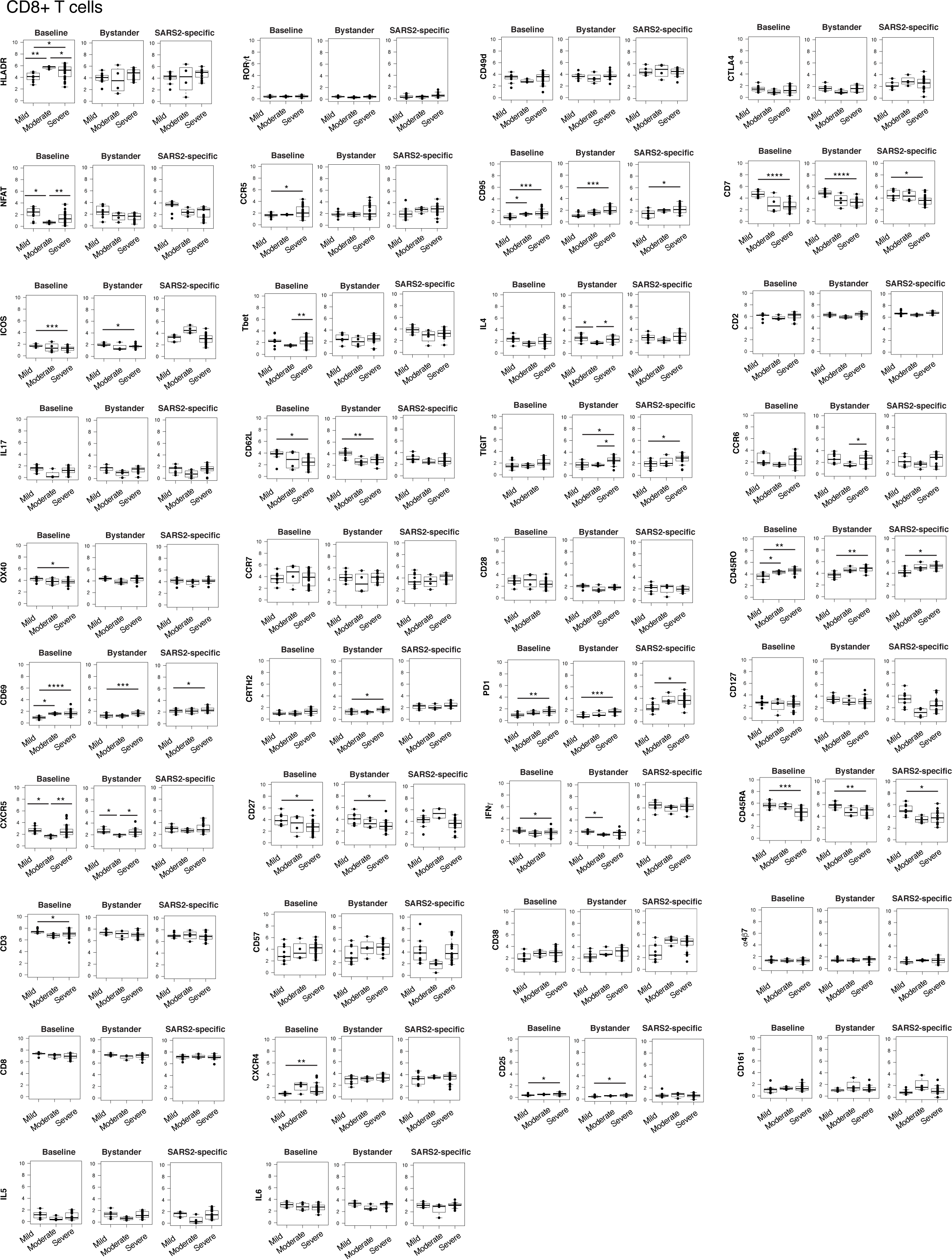
Mean signal intensity reflecting expression levels of 38 surface and intracellular antigens in baseline, bystander, and SARS-CoV-2-specific CD8+ T cells from mild, moderate, and severe COVID-19 – Related to Figure 1. PBMCs were purified from freshly drawn blood specimens and phenotyped by CyTOF at baseline or following a 6-hour stimulation with anti-CD49d/CD28 and overlapping 15-mer peptides from SARS-CoV-2 spike. Bystander cells are cells that remained IFNγ-in the stimulated samples. SARS-CoV-2-specific CD8+ T cells are the cells that produced IFNγ following stimulation. Shown are the mean signal intensity (MSI) values for arcsinh-transformed data. *p < 0.05, **p < 0.01, ***p < 0.001, ****p < 0.0001 as assessed using the Student’s unpaired t-test and adjusted for multiple testing using the Benjamini-Hochberg for FDR.

**Figure S4.**
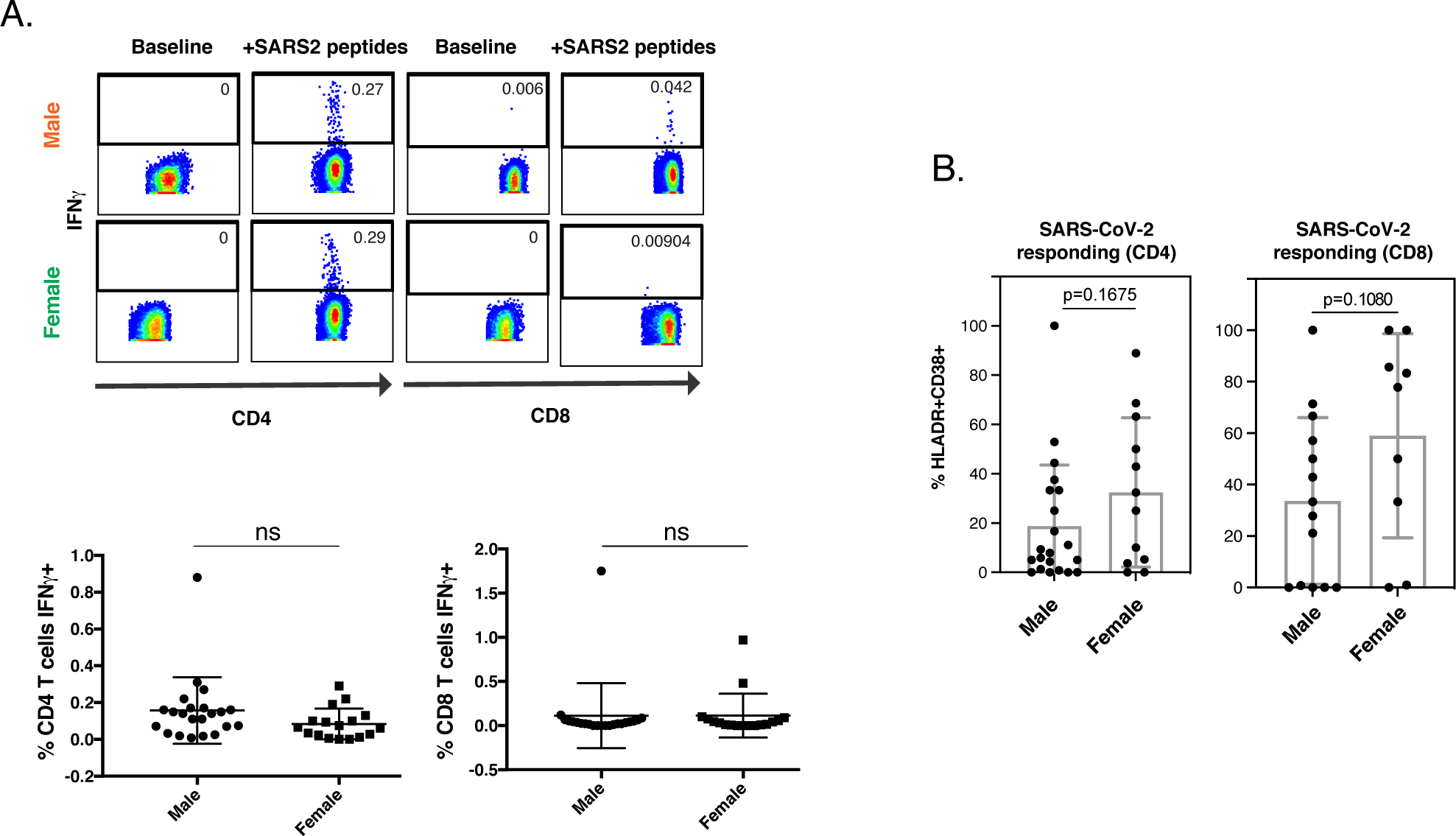
Male and female hospitalized COVID-19 patients harbor similar frequencies of SARS-CoV-2-specific T cells – Related to Figure 5. (**A**) SARS-CoV-2-specific T cells in male and female patients hospitalized for COVID-19. *Top:* Representative pseudocolor plots of CyTOF datasets showing the percentage of SARS-CoV-2-specific CD4+ or CD8+ T cells from infected male or female patients in the ICU. *Bottom:* Graphs showing the cumulative data from all infected hospitalized individuals. ns: non-significant as determined by a Student’s unpaired t-test. (**B**) SARS-CoV-2-specific CD4+ and CD8+ T cells from hospitalized patients tend to be more activated (HLADR+CD38+) in females than in males. p-values were calculated using the Student’s unpaired t-test.

**Figure S5.**
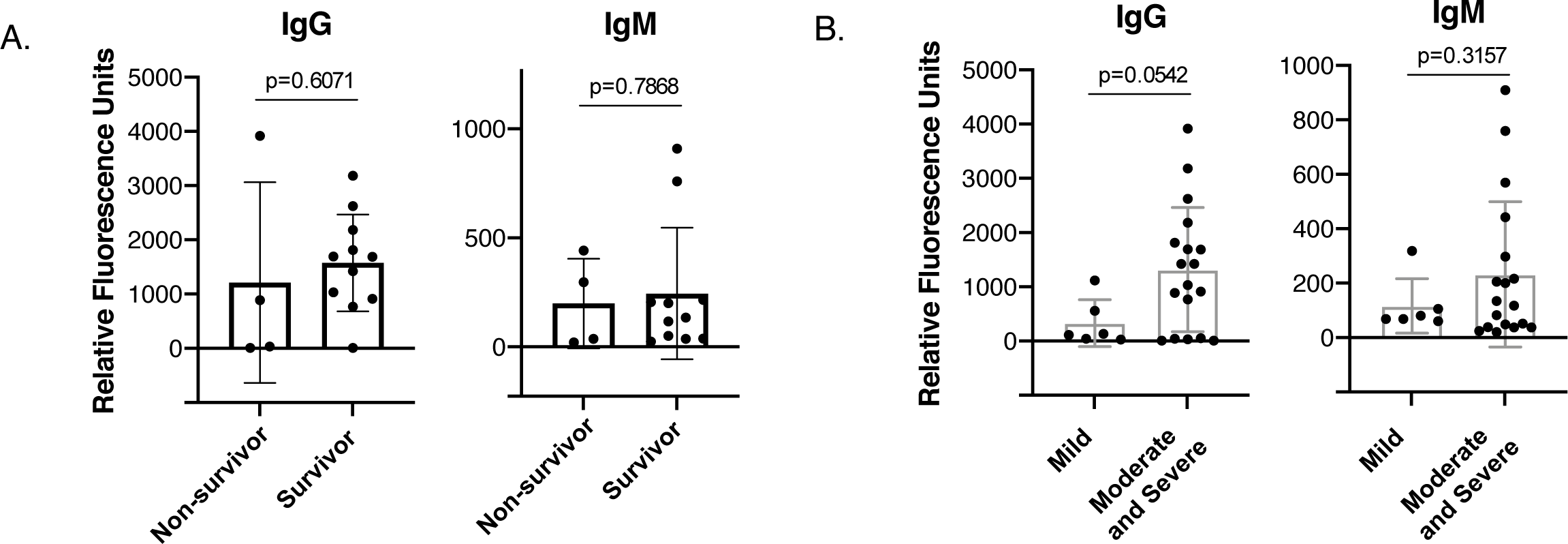
Individuals that succumb to severe COVID-19 do not harbor higher levels of anti-SARS-CoV-2 antibodies than those that survive – Related to Figure 5. (**A**) Anti-SARS-CoV-2 antibody levels are similar between non-surviving and surviving patients hospitalized in the ICU for COVID-19. The levels of IgG and IgM against the spike RBD and nucleocapsid of SARS-CoV-2 were measured using an immunoassay system. (**B**) Anti-SARS-CoV-2 antibody levels exhibit tend to be elevated in hospitalized patients (moderate and severe groups) relative to outpatients (mild group). Only the subset of patients that did not receive convalescent plasma (Table S1) were used in this analysis.

**Figure S6.**
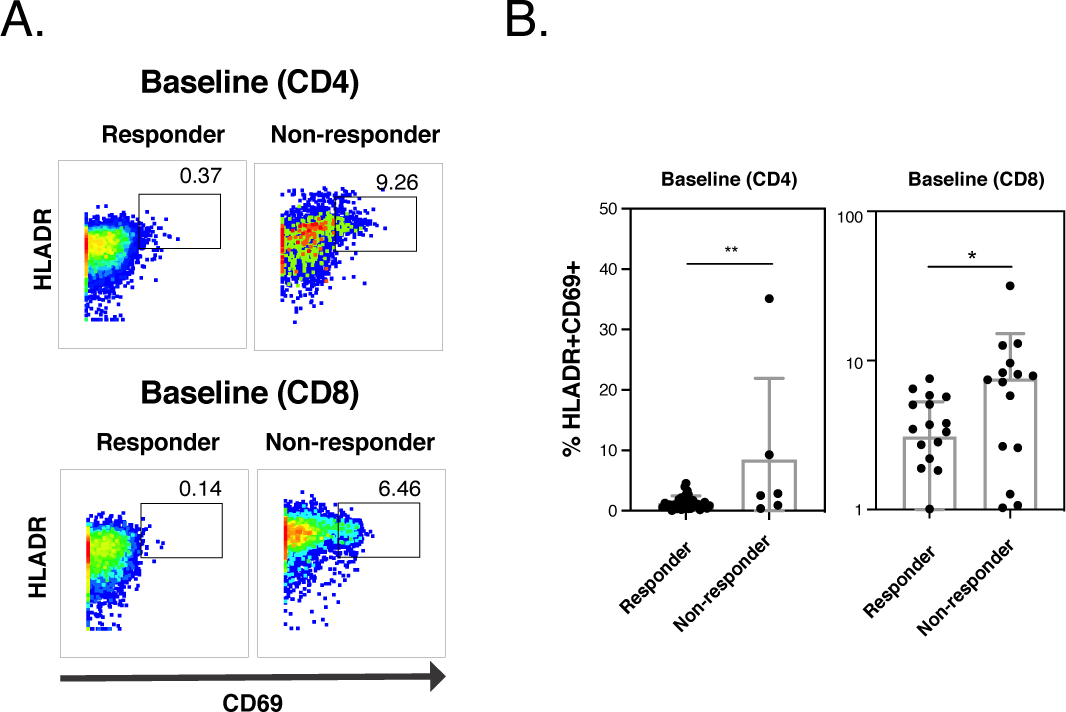
Hospitalized COVID-19 patients that mounted SARS-CoV-2-specific T cell responses harbor fewer total activated T cells – Related to Figure 5. (**A**) Pseudocolor dot plots from representative hospitalized patients that mounted (“responder”) vs. did not mount (“non-responder”) SARS-CoV-2-specific T cell responses. The percentages of activated (HLADR+CD69+) cells are indicated for total CD4+ (*top*) or CD8+ (*bottom*) T cells. (**B**) Graphic representation of the data shown in (*A*) across all responder and non-responder hospitalized patients. * p < 0.05 and * p < 0.01 as determined by a Student’s unpaired t-test.

**Figure S7.**
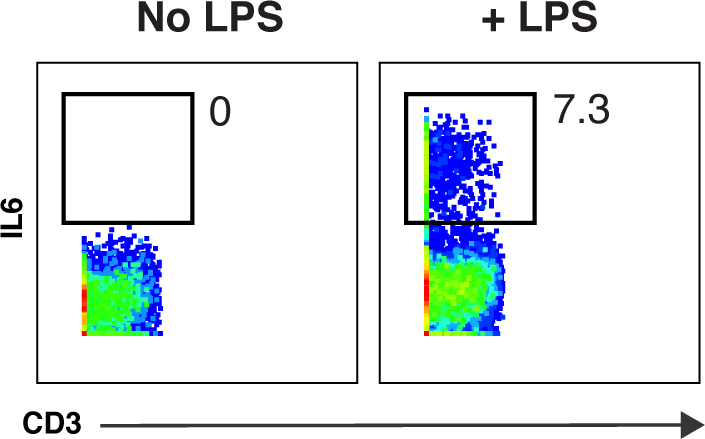
Detection of robust IL6 production by CyTOF upon stimulation of monocytes with LPS – Related to Figure 5. Peripheral blood monocytes (CD3-CD4^dim^ cells) were stimulated with LPS in the presence of brefeldin A for 6 hours, and then analyzed by CyTOF. The detection of IL6 in the LPS-stimulated but not in the unstimulated sample demonstrates the specificity and sensitivity of the IL6 antibody used for CyTOF analysis.

**Figure S8.**
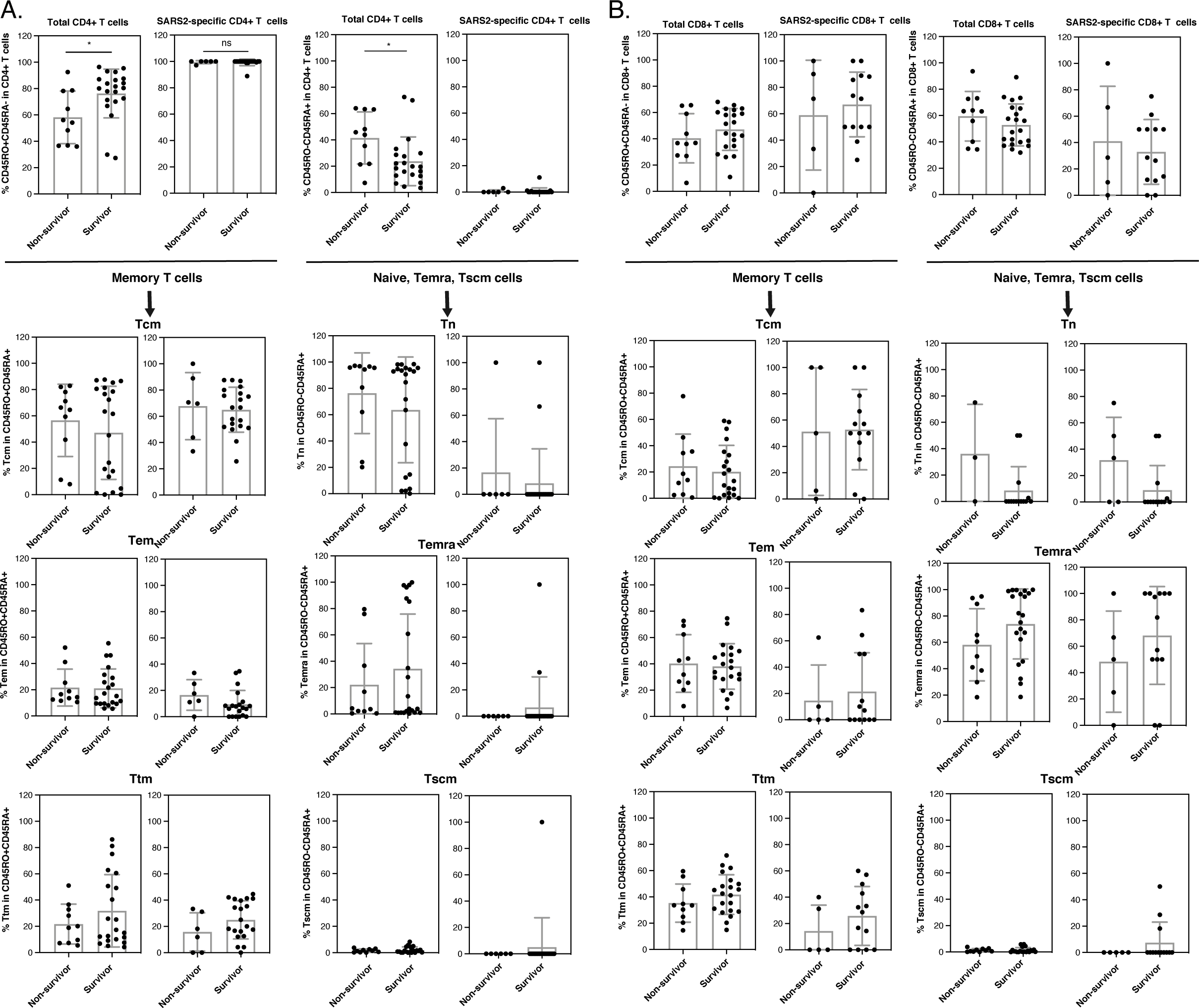
Subset analysis of total and SARS-CoV-2-specific T cells from fatal vs. non-fatal cases of severe COVID-19 – Related to Figure 5. The subsets described in Fig. 2 were compared between infected ICU patients that did or did not survive severe infection. A significant increase in memory cells among the total CD4+ T cell compartment was observed in the survivors, and this was accompanied by a significant decrease in CD45RO-CD45RA+ cells, which includes Tn cells. * p < 0.05 as determined by a Student’s unpaired t-test.

**Figure S9.**
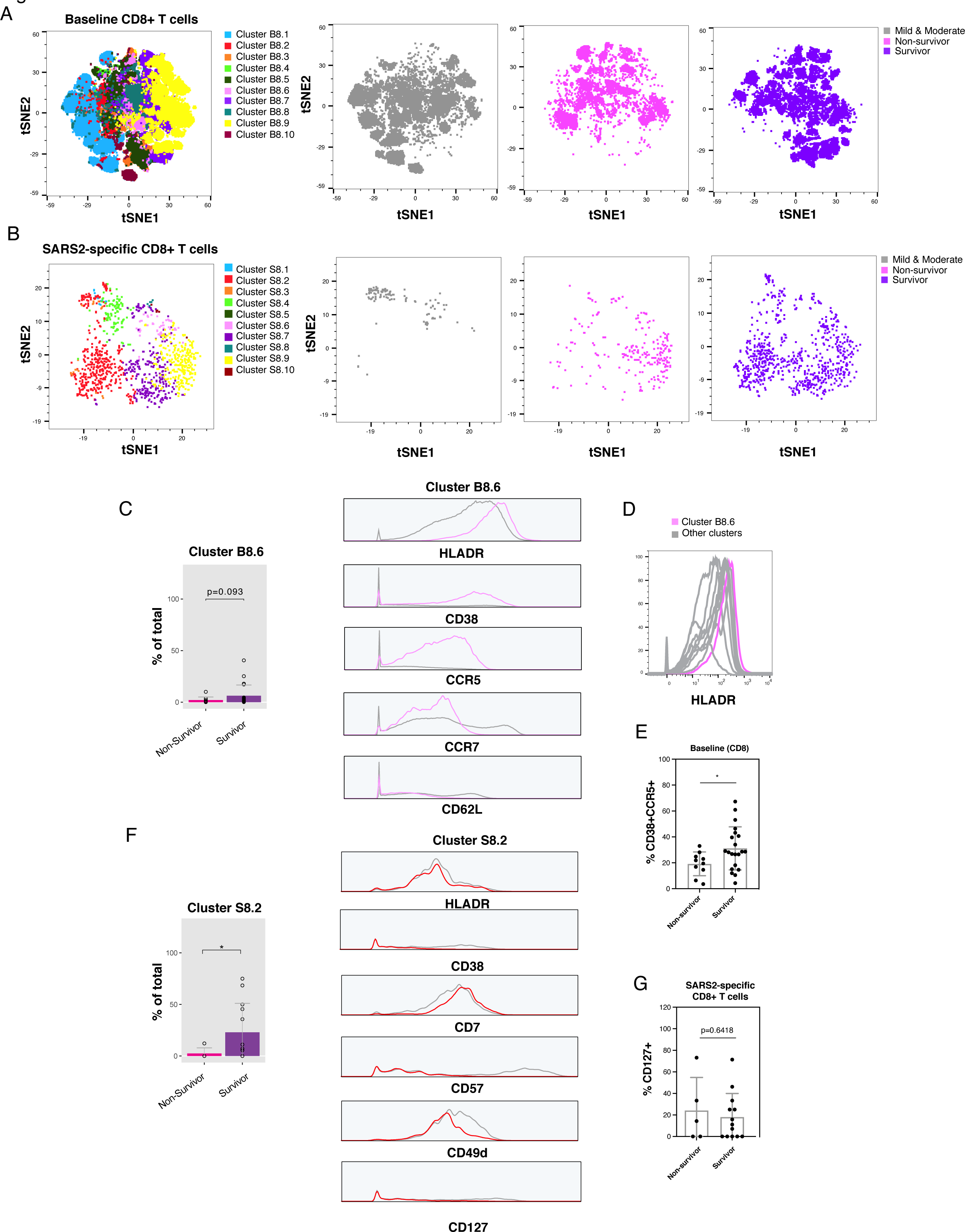
Clustering analysis reveals enrichment of activated total and non-terminally differentiated SARS-CoV-2-specific CD8+ T cells in patients that recover from severe COVID-19 – Related to. Figure 6. (**A**) The phenotypes of total CD8+ T cells differ in non-survivors vs. survivors of severe COVID-19. Shown on the left are the FlowSOM clusters identified in Fig. 4A for reference. The 3 plots on the right show the distribution of cells among non-survivors and survivors visualized by t-SNE, relative to the other patient groups (mild and moderate). (**B**) The phenotypes of SARS-CoV-2-specific CD8+ T cells differ in non-survivors vs. survivors of severe COVID-19. The analyses described in *panel A* were conducted among SARS-CoV-2-specific CD8+ T cells. (**C**) Activated memory CD8+ T cells are increased in survivors of severe COVID-19. *Left:* Cluster C8.6 of total CD8+ T cells was enriched in the survivor relative to the non-survivor group. *Right:* Cells from cluster B8.6 express high levels of activation markers HLADR, CD38, and CCR5, and low levels of the Tcm markers CCR7 and CD62L. Expression levels from cluster B8.6 are displayed in pink, while those from total CD8+ T cells are displayed in grey. (**D**) Among all clusters of total CD8+ T cells, cells from cluster B8.6 express the highest levels of HLADR. (**E**) Survivors of severe COVID-19 express elevated frequencies of activated (CD38+CCR5+) CD8+ T cells. (**F**) Survivors of severe COVID-19 harbor more SARS-CoV-2-specific CD8+ T cells expressing low levels CD57 and CD49d. *Left:* Cluster S8.2 was enriched in survivors. *Right:* Cells from cluster S8.2 do not express elevated levels of HLADR, CD38, or CD127, and express low levels of the terminal differentiation marker CD57 and the integrin component CD49d. Expression levels from cluster S8.2 are displayed in pink, while those from total SARS-CoV-2 CD8+ T cells are displayed in grey. * p < 0.05 as determined by a Student’s unpaired t-test. (**G**) The proportions of CD127+ cells among SARS-CoV-2-specific CD8+ T cells are similar between survivors and non-survivors of severe COVID-19. * p < 0.05 as determined by a Student’s unpaired t-test.

**Figure S10.**
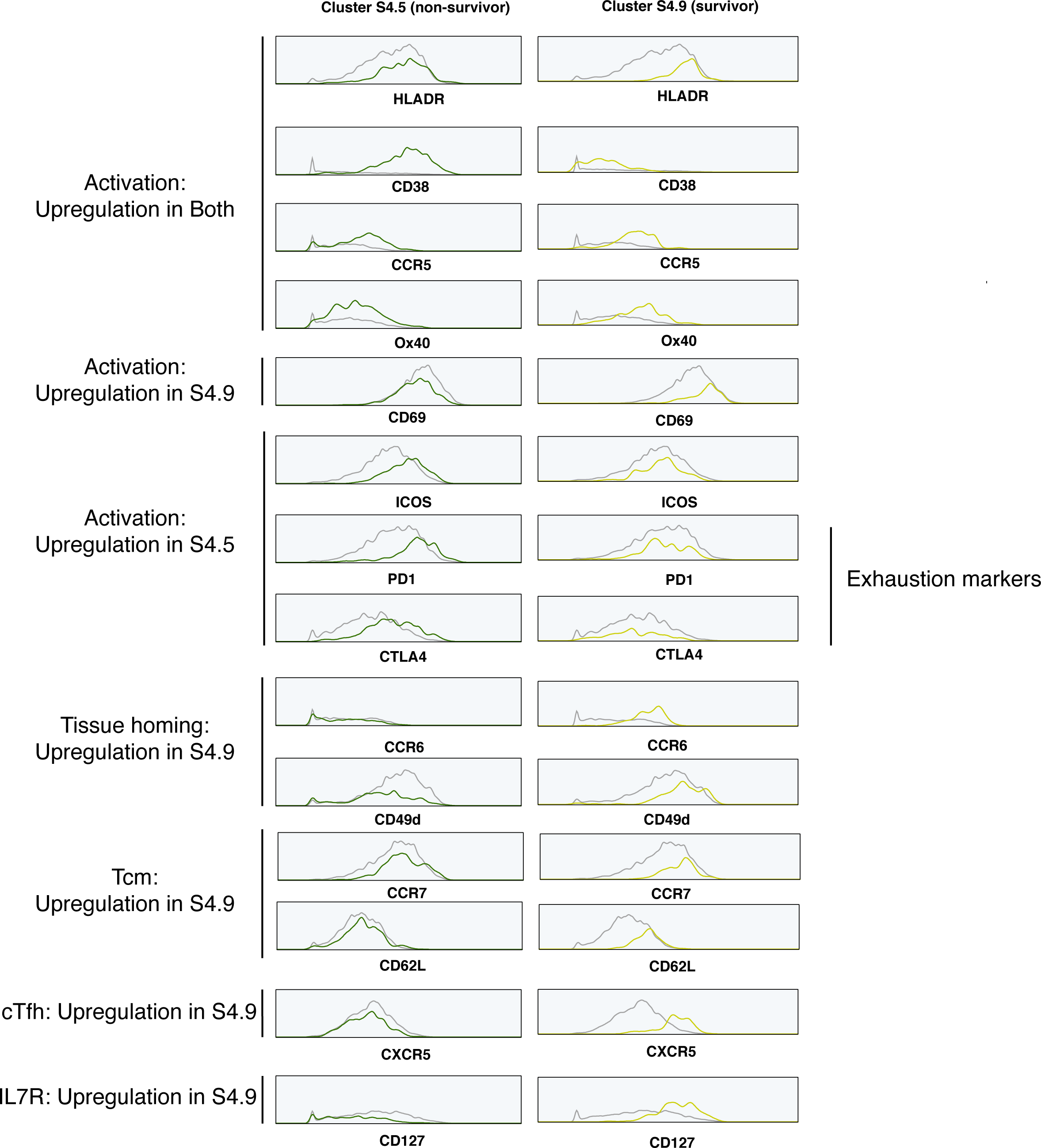
Distinct phenotypes of SARS-CoV-2-specific CD4+ T cell clusters enriched in survivors vs. non-survivors of severe COVID-19 – Related to Figure 6. SARS-CoV-2-specific FlowSOM cluster S4.5 (enriched in non-survivors) and S4.9 (enriched in survivors) described in Fig. 6E were examined in more detail. Among activation markers, both clusters expressed elevated levels of HLADR, CD38, CCR5, and Ox40, while S4.9 selectively upregulated CD69, and S4.5 selectively upregulated ICOS, PD1, and CTLA4. Of note, PD1 and CTLA4 additionally serve as exhaustion markers. The tissue homing receptors CCR6 and CD49d, Tcm markers (CCR7, CD62L), the cTfh marker CXCR5, and CD127 were upregulated in cluster S4.9.

**Figure S11.**
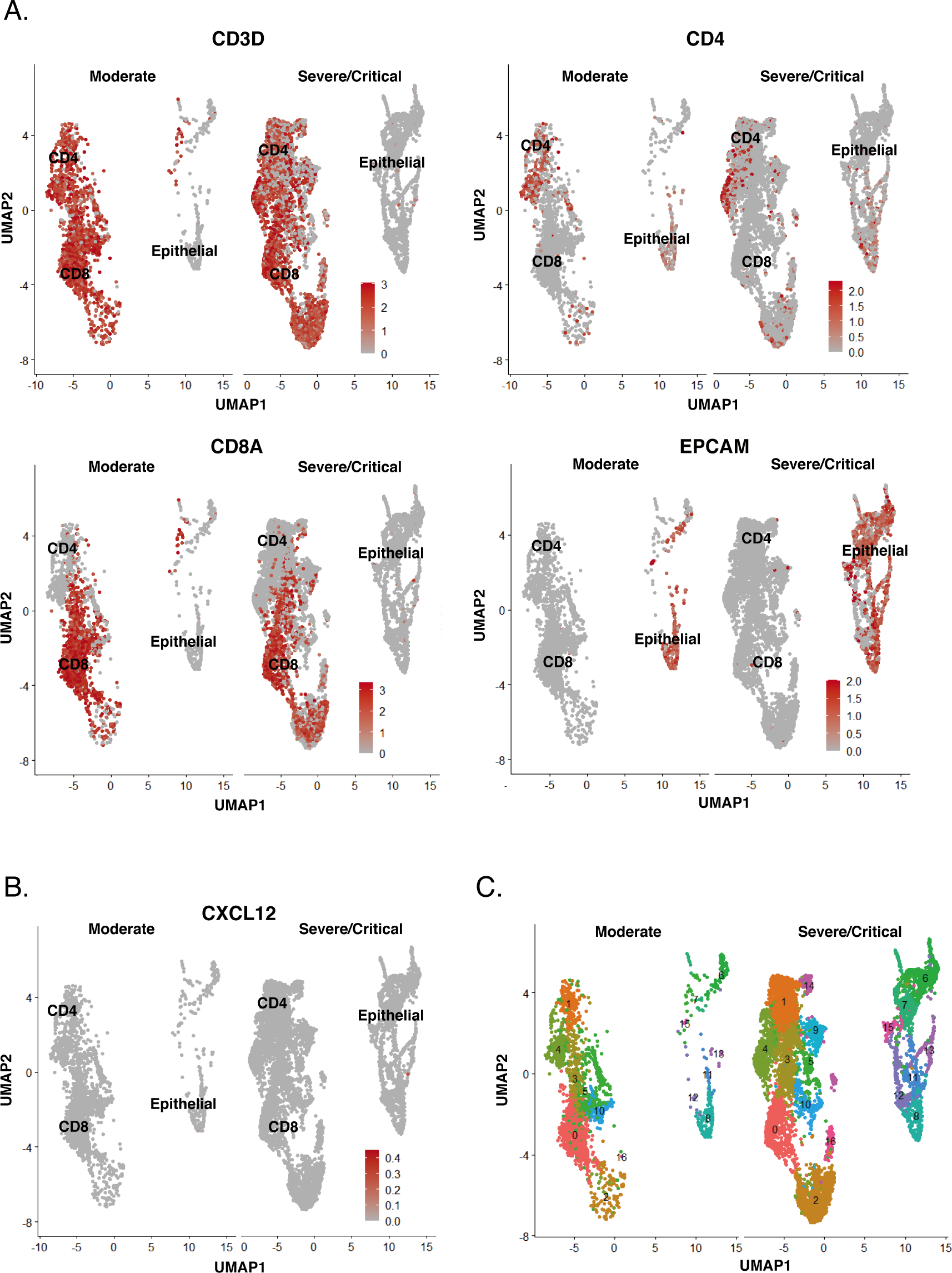
Subset identification of T and epithelial cells from lungs of COVID-19 patients – Related to Figure 7. (**A**) The CD4+ T cell, CD8+ T cell, and epithelial cell subsets were extracted from public scRNAseq datasets generated from BAL of COVID-19 patients with moderate or severe/critical disease as illustrated in Fig. 7F, and verified by assessing RNA expression levels of the T cell marker CD3D, CD4, the alpha chain of CD8 (CD8A), and the epithelial marker EpCAM. Expression levels are depicted as UMAP heatmaps. (**B**) In contrast to HMGB1 (Fig. 7I), the CXCR4 ligand CXCL12 is poorly expressed in T and epithelial cells of the lung. (**C**) Clustering of pulmonary T and epithelial cells from COVID-19 patients reveals CD8+ T cells with highest levels of HMGB1 expression (Fig. 7I) to form a unique cluster, cluster 2.

**Table S1.** COVID-19 Patient Characteristics

| <u>Patient ID*</u> | <u>Gender</u> | <u>Age Range</u> | <u>Race</u> | <u>Severity</u> | <u>Conva-lescent Plasma</u> | <u>Dexa-methasone</u> | <u>Rem-desivir</u> | <u>Days from discharge or death*</u> | <u>Days between 1<sup>st</sup> PCR test and analysis*</u> |
| --- | --- | --- | --- | --- | --- | --- | --- | --- | --- |
| COV26 | Female | 80-84 | LatinX | Moderate | No | No | Yes | 8 | 2 |
| COV21 | Female | 45-50 | LatinX | Moderate | No | No | Yes | 1 | 6 |
| COV16 | Female | 45-50 | AA | Moderate | No | No | No | 0 | 13 |
| COV25 | Male | 50-54 | LatinX | Moderate | No | No | No | 20 | 1 |
| COV06 | Male | 50-54 | LatinX | Moderate | No | No | Yes<sup>1</sup> | 3 | 8 |
| COV07 | Male | 60-64 | Asian | Moderate | Yes | No | Yes | 5 | 11 |
| COV14 | Male | 45-49 | Asian | Severe | No | Yes | Yes | 8 | 15 |
| COV05 | Male | 70-75 | LatinX | Severe | Yes | No | No | 34 | 28 |
| COV13 | Female | 60-64 | AA | Severe | No | No | Yes | 11 | 10 |
| COV42 | Female | 70-74 | LatinX | Severe | No | Yes | Yes | 8 | 8 |
| COV47 | Female | 65-69 | AA | Severe | No | Yes | Yes | 25 | 0 |
| COV61 | Female | 65-69 | Asian | Severe | No | No | Yes | 5 | 14 |
| COV04 | Male | 75-79 | Asian | Severe | No | No | No | 4 | 54 |
| COV80 | Female | 55-59 | LatinX | Severe | No | Yes | Yes | 1 | 26 |
| COV08<br>COV12<br>COV17 | Male | 45-49 | LatinX | Severe | No | No | No | 56<br>30<br>16 | 1<br>27<br>41 |
| COV03<br>COV11 | Male | 60-64 | LatinX | Severe | No | No | Yes | 35<br>16 | 57<br>76 |
| COV19<br>COV37 | Male | 65-69 | LatinX | Severe | Yes | No | Yes | NA<br>NA | 32<br>42 |
| COV20<br>COV38<br>COV53b<br>COV69 | Female | 55-59 | LatinX | Severe | Yes | Yes | Yes | 56<br>46<br>32<br>28 | 10<br>20<br>34<br>38 |
| COV31<br>COV46<br>COV74<br>COV86 | Male | 60-64 | LatinX | Severe | Yes | No | No | 45<br>43<br>25<br>17 | 7<br>9<br>27<br>35 |
| COV01 | Female | 30-34 | LatinX | Severe/Death | No | No | Yes | 35 | 14 |
| COV18 | Female | 75-79 | LatinX | Severe/Death | Yes | Yes | Yes | 12 | 10 |
| COV15 | Female | 80-84 | LatinX | Severe/Death | No | No | Yes | 8 | 9 |
| COV02<br>COV10 | Female | 80-84 | LatinX | Severe/Death | No | No | Yes | 19<br>0 | 2<br>21 |
| COV22<br>COV24<br>COV39 | Male | 65-96 | LatinX | Severe/Death | Yes | Yes | Yes | 39<br>13<br>11 | 5<br>13<br>15 |
| COV45<br>COV62 | Male | 70-74 | LatinX | Severe/Death | Yes | No | No | 14<br>3 | 7<br>21 |
| PID4123 | Male | 30-34 | White | Mild | No | No | No | NA | 25 |
| PID4103 | Female | 40-44 | White | Mild | No | No | No | NA | 20 |
| PID4100 | Female | 25=29 | White | Mild | No | No | No | NA | 36 |
| PID4114 | Female | 45-49 | White | Mild | No | No | No | NA | 59 |
| PID4104 | Female | 30-34 | White | Mild | No | No | No | NA | 60 |
| PID4122 | Female | 30-34 | White | Mild | No | No | No | NA | 26 |
| PID4106 | Female | 65-69 | White | Mild | No | No | No | NA | 46 |
| PID4112 | Female | 55-59 | White | Mild | No | No | No | NA | 154 |
| PID4102 | Male | 45-49 | White | Mild | No | No | No | NA | 47 |

\* Multiple entries = multiple timepoints from same patient available
Yes<sup>1</sup> = enrolled in a placebo-controlled remdesivir trial
**Race:** LatinX = Hispanic or Latino, AA = Black or African-American
**Severity:** Severe = ICU, Moderate = Admitted, Death = Death by COVID-19, Mild = Outpatient convalescent
**NA:** Not Available or Not Available

**Table S2.** List of CyTOF antibodies used in study. Antibodies were either purchased from the indicated vendor or prepared in-house using commercially available MaxPAR conjugation kits per manufacturer’s instructions (Fluidigm).

| **Antigen Target** | **Clone** | **Elemental Isotope** | **Vendor** |
| --- | --- | --- | --- |
| HLADR | TÜ36 | Qdot (112Cd) | Thermofisher |
| RORγt* | AFKJS-9 | 115 In | In-house |
| CD49d (a4) | 9F10 | 141Pr | Fluidigm |
| CTLA4* | 14D3 | 142Nd | In-house |
| NFAT* | D43B1 | 143Nd | Fluidigm |
| CCR5 | NP6G4 | 144Nd | Fluidigm |
| IL5* | TRFK5 | 145Nd | In-house |
| CD95 | BX2 | 146Nd | In-house |
| CD7 | CD76B7 | 147Sm | Fluidigm |
| ICOS | C398.4A | 148Nd | Fluidigm |
| Tbet* | 4B10 | 149Sm | In-house |
| IL4* | MP4-25D2 | 150Nd | In-house |
| CD2 | TS1/8 | 151Eu | Fluidigm |
| IL17* | BL168 | 152Sm | In-house |
| CD62L | DREG56 | 153Eu | Fluidigm |
| TIGIT | MBSA43 | 154Sm | Fluidigm |
| CCR6 | 11A9 | 155Gd | In-house |
| IL6* | MQ2-13A5 | 156 Gd | In-house |
| CD8 | RPA-T8 | 157Gd | In-house |
| CD19 | HIB19 | 157Gd | In-house |
| CD14 | M5E2 | 157Gd | In-house |
| OX40 | ACT35 | 158Gd | Fluidigm |
| CCR7 | G043H7 | 159Tb | Fluidigm |
| CD28 | CD28.2 | 160Gd | Fluidigm |
| CD45RO | UCHL1 | 161Dy | In-house |
| CD69 | FN50 | 162Dy | Fluidigm |
| CRTH2 | BM16 | 163Dy | Fluidigm |
| PD-1 | EH12.1 | 164Dy | In-house |
| CD127 | A019D5 | 165Ho | Fluidigm |
| CXCR5 | RF8B2 | 166Er | In-house |
| CD27 | L128 | 167Er | Fluidigm |
| IFNγ* | B27 | 168Er | Fluidigm |
| CD45RA | HI100 | 169Tm | Fluidigm |
| CD3 | UCHT1 | 170Er | Fluidigm |
| CD57 | HNK-1 | 171Yb | In-house |
| CD38 | HIT2 | 172Yb | Fluidigm |
| α4β7 | Act1 | 173Yb | In-house |
| CD4 | SK3 | 174Yb | Fluidigm |
| CXCR4 | 12G5 | 175Lu | Fluidigm |
| CD25 | M-A251 | 176Yb | In-house |
| CD161 | NKR-P1A | 209 Bi | In-house |

\*Intracellular antibodies

## Notes

### Competing Interest Statement

The authors have declared no competing interest.

### Author Declarations

This study was approved by the University of California, San Francisco (UCSF) Human Research Protection Program (HRPP). The institutional review board (IRB) approval number is IRB # 20-30588.

